# Data Extraction for Evidence Synthesis Using a Large Language Model: A Proof-of-Concept Study

**DOI:** 10.1101/2023.10.02.23296415

**Authors:** G Gartlehner, L Kahwati, R Hilscher, I Thomas, S Kugley, K Crotty, M Viswanathan, B Nussbaumer-Streit, G Booth, N Erskine, A Konet, R Chew

**Affiliations:** Social, Statistical, and Environmental Sciences, RTI International, Research Triangle Park, USA; Department for Evidence-based Medicine and Evaluation, Danube University Krems, Krems, Austria; Preventive Medicine Residency Program, Department of Family Medicine, School of Medicine, University of North Carolina at Chapel Hill, Chapel Hill, USA

## Abstract

Data extraction is a crucial, yet labor-intensive and error-prone part of evidence synthesis. To date, efforts to harness machine learning for enhancing efficiency of the data extraction process have fallen short of achieving sufficient accuracy and usability. With the advent of Large Language Models (LLMs), new possibilities have emerged to increase efficiency and accuracy of data extraction for evidence synthesis. The objective of this proof-of-concept study was to assess the performance of an LLM (Claude 2) in extracting data elements from published studies, compared with human data extraction as employed in systematic reviews. Our analysis utilized a convenience sample of 10 English-language, open-access publications of randomized controlled trials included in a single systematic review. We selected 16 distinct types of data, posing varying degrees of difficulty (160 data elements across 10 studies). We used the browser version of Claude 2 to upload the portable document format of each publication and then prompted the model for each data element. Across 160 data elements, Claude 2 demonstrated an overall accuracy of 96.3% with a high test-retest reliability (replication 1: 96.9%; replication 2: 95.0% accuracy). Overall, Claude 2 made 6 errors on 160 data items. The most common errors (n=4) were missed data items. Importantly, Claude 2’s ease of use was high; it required no technical expertise or training data for effective operation. Based on findings of our proof-of-concept study, leveraging LLMs has the potential to substantially enhance the efficiency and accuracy of data extraction for evidence syntheses.

## Background

Systematic reviews (SRs) and other types of evidence syntheses are the benchmark for assessing the efficacy and risks of healthcare interventions, treatments, diagnostic tests, and technologies.^1^ Conducting evidence synthesis involves standardized steps, such as formulating precise research questions, conducting comprehensive literature searches, critically appraising the methods of eligible studies, extracting data from included studies, and synthesizing evidence.^2^ Among these, data extraction from selected studies (i.e., the process of manually extracting data from primary studies into standardized tables) is one of the most time-consuming, costly, and crucial tasks in evidence synthesis.^3^ In a randomized trial assessing different data extraction strategies, single investigator data extraction and verification by a second investigator took, on average, 107 minutes per study, dual independent data extraction took 172 minutes.^4^ Data extraction errors can seriously undermine the validity of evidence syntheses, as they can affect narrative summaries, meta-analyses, and conclusions. A methodological review revealed a high rate of data extraction errors (up to 63%) in systematic reviews.^5^ The error rate varied depending on the type and complexity of the data.^5^ The causes of data extraction errors are multifaceted, including inaccuracies such as missing available data, misclassifications (e.g., mistaking a standard deviation for a standard error), misinterpretations stemming from ambiguous reporting in primary studies, or straightforward data entry mistakes. Factors such as time constraints, and language barriers can further heighten the risk of data extraction errors.^6-8^

The use of artificial intelligence (AI) can potentially increase efficiency of the data extraction process. Semi-automation refers to the partial automation of certain tasks in the data extraction process while retaining human involvement. For example, natural language processing (NLP) algorithms can help extract specific information, such as study characteristics, outcomes, or effect estimates from the full-text articles. However, human reviewers may still need to validate and cross-check the extracted data to ensure accuracy, completeness, and consistency. Research on methods for semi-automating data extraction in the past has mostly focused on NLP using statistical models such as naïve Bayes or support vector machines.^9^ All of these models require training data and often encounter difficulties in extracting information from articles in portable document format (PDF), especially tables. In general, the training of NLP models to extract data is both time-consuming and resource intensive. A living systematic review on automated and semi-automated data extraction methods found 53 publications since 2005.^9^ Most studies addressed data extraction from abstracts alone; only eight addressed extraction from full-text articles in the form of PDFs. The findings from research on this topic suggest that tools for automated or semi-automated data extraction are still not mature enough for practical use.^7^

With the advent of notable commercial Large Language Models (LLMs), such as Generative Pre-trained Transformer-4 (GPT-4)^10^ and Claude 2^11^, new possibilities have emerged to increase efficiency of data extraction if these new AI technologies can be adapted for use in evidence synthesis. An LLM is a type of machine learning model specifically designed to predict, generate, and comprehend human-like text.^12^ Generative LLMs are trained to perform a “language modeling” task, in which the objective is to predict the next token, conditional on a prior sequence of text. This flexible objective allows LLMs to be multi-purpose; by structuring the input text as a set of instructions (i.e., “prompts”), they are capable of performing a wide variety of text generation and comprehension tasks.^13^ However, an LLMs capabilities can vary dramatically based on aspects such as the number of model parameters^14^ and the kinds of fine-tuning performed.^15,16^ At the time of writing, commercial LLMs created by AI research labs, like those evaluated in this work, tend to greatly outperform open-source models.^17^

The primary objective of this proof-of-concept study was to assess the performance of an LLM in extracting pre-specified data elements from PDF versions of full-text study reports published in scientific journals (henceforth referred to as “study reports”) compared to data extraction by humans as employed in SRs. The study aimed to investigate the preliminary accuracy, and reliability of data extraction capabilities for data elements that are commonly used for SRs or other evidence syntheses. Table 1 provides definitions of commonly used terms in this manuscript.

**Table 1.**
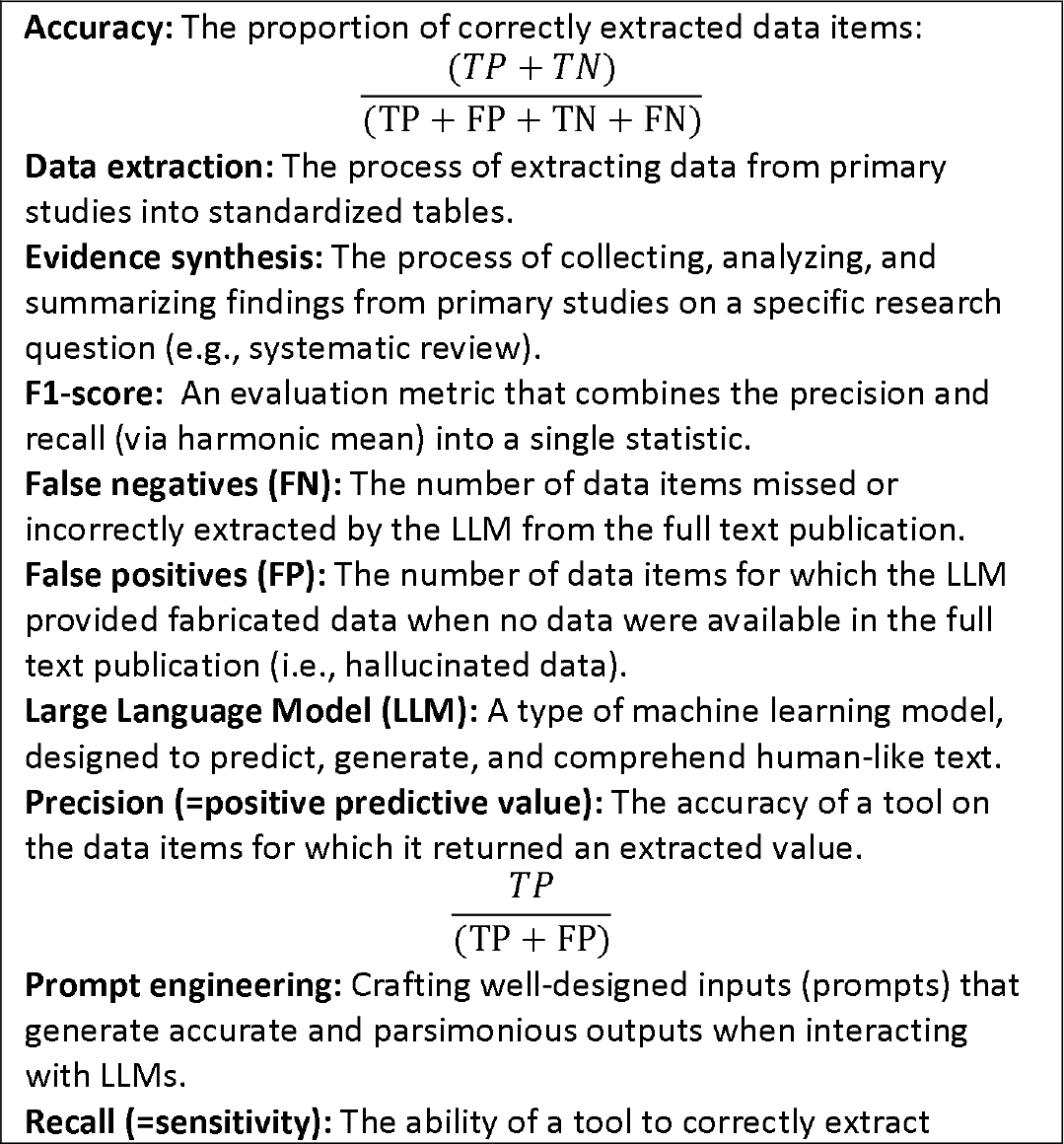
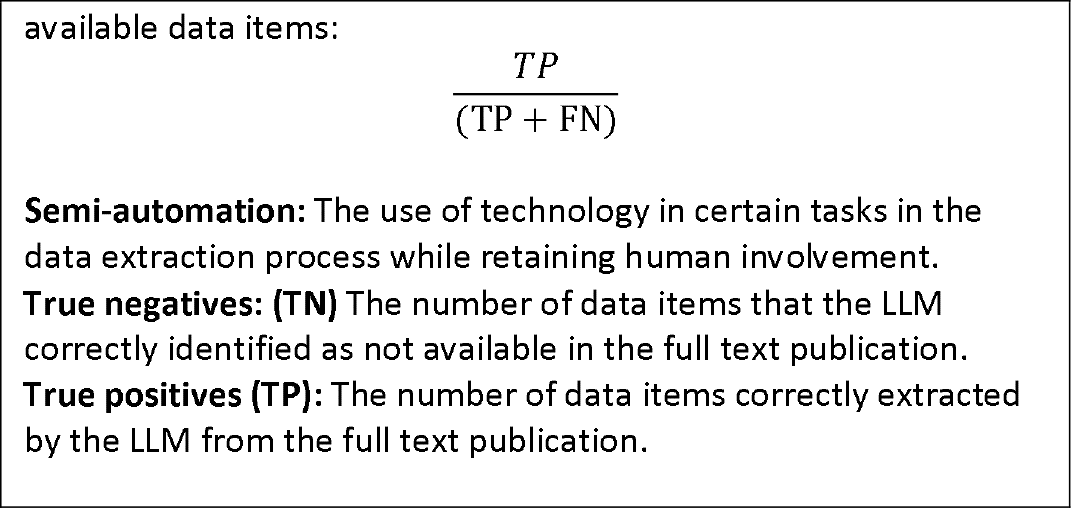
Definitions of Commonly Used Terms.

## Methods

We registered the protocol of this study in the Open Science Framework: osf.io/2546n. We had originally intended to only evaluate the GPT-4 LLM, but on August 4, 2023, we amended our protocol to use Claude 2 instead of GPT-4 because Claude 2 natively supports the direct upload of PDF documents and has a large enough context window (100,000 tokens [i.e., individual pieces of a text such as words, characters, or sub-words]) to include an entire article as part of a prompt.

### Study design

We employed a validation study design that compared the performance of Claude 2 with a reference standard to address the following research questions:

1. How does the accuracy of data extraction from PDF versions of study reports using Claude 2 compare to dual manual extraction methods by humans, as employed in SRs?
2. What is the reliability and consistency of data extraction by Claude 2 across multiple studies on the same topic?
3. What type of errors does Claude 2 make when extracting data from study reports?
4. Which types of data are most likely to be extracted accurately by Claude 2?

#### Selection of reference standard

We used data previously extracted by a single investigator and reviewed for accuracy against the source PDF document by a second investigator from a convenience sample of 10 English-language open-access study reports^18-27^of randomized controlled trials (RCTs), included in a previously conducted systematic review on targeted immune modulators for the treatment of plaque psoriasis. We selected publications of RCTs of medications due to their well-defined study design and standardized reporting compared to other study types. Because this was a proof-of-concept study, we did not perform sample size calculations.

We selected 16 data elements representing four distinct types of data (e.g., numeric, text) that pose varying degrees of difficulty for the extraction process: 1) Study identifiers (e.g., registration number, first author); 2) Characteristics of study participants (e.g., mean age, inclusion/exclusion criteria); 3) Numerical data related to participant flow (e.g., number of randomized individuals overall and per study group); and 4) Primary outcome specified and outcome data (e.g., name of outcome, the proportion of individuals experiencing the outcome). For this study, we focused on dichotomous outcome data.

#### Prompt Engineering

Prompt engineering^13^ involves crafting well-designed instructions (prompts) to generate accurate and parsimonious outputs when interacting with LLMs such as Claude 2. The initial prompts were carefully crafted, relying on a clear definition of each data element. We then conducted iterative testing on three of the 10 articles included for this study to develop effective prompts for each data element. When required, we steered the model towards a preferred response format, such as specifying the number of decimal places. We refined the prompts if data extraction was incomplete, or if the output was in an unsuitable format. Appendix 1 presents the final prompts for each data element.

#### Data Extraction and Analysis

We used the browser version of Claude 2 to upload the PDF of each study report and then prompted the model for each data element. We then compared data extracted by Claude 2 with the reference standard. When discrepancies between the LLM and reference standard occurred, we checked the respective full-text source PDF to validate the accuracy of the reference standard. Because human-led data extraction is an imperfect reference standard, we followed guidance by the Agency for Healthcare Research and Quality^28^ and made necessary corrections if errors in the reference standard were identified. When prompt revisions were necessary, we used data extracted for the final prompt for our analyses.

To better understand and classify erroneous data extractions by Claude 2, we developed a classification system of four types of errors (Table 2). One investigator classified the types of errors, a second investigator reviewed classifications for correctness.

**Table 2:** Types of errors for data extraction with large language models.

| Types of error | Definitions |
| --- | --- |
| Major error | This error significantly compromises the accuracy of the data, and, if uncorrected, could lead to erroneous conclusions; for example, grossly incorrect calculations or misallocated data. |
| Minor error | This error is less severe than a major error but still impacts the quality of the |
|  | existing data; for example, small calculation errors or rounding errors that do not critically affect the data's overall utility. |
| False data ("Hallucination") | Fabricated data that seem to be generated by the LLM |
| Missed or omitted data | Data that were available in the reference standard (or source PDF) but were either missed or omitted by the LLM. |

Abbreviations: LLM = large language model

We aggregated the results in a contingency table and calculated global accuracy estimates (accuracy and F1 score). Our unit of analysis consisted of individual data items. Therefore, if multiple errors occurred within the same data item—such as an incorrect mean and a misreported standard deviation for mean age—we counted them as a single error.

To evaluate test-retest reliability, we employed the final prompts in two replications, using the same sample articles four weeks after the initial data extractions.

All statistical analyses were conducted with Stata 16.1 (StataCorp LLC, Texas, USA).

#### Data Management

The study used open-access scientific publications. We stored data electronically in Excel datasheets. All investigators had access to the data.

## Results

When we reviewed initial discrepancies between data extracted by Claude 2 and the reference standard, we identified 21 instances of minor errors in the reference standard data (detailed in Appendix 2). Examples included incorrectly transcribed numeric values, rounding errors, minor errors in dosing intervals, and missing exclusion criteria or baseline characteristics. In these cases, we corrected the reference standard data based on the source PDF publication prior to comparing with data extracted by Claude 2. In addition, we realized that in two instances, the reference standard contained additional detail related to inclusion or exclusion criteria that human extractors had obtained from companion publications (i.e., related publications reporting on the same study as the source PDF). We did not count these instances as data omissions by Claude 2, as we did not provide Claude 2 with these companion publications for analysis.

### Accuracy of data extractions

Out of a total of 160 data elements across 10 study reports, complete information was reported in the reference standard for 157 items. Overall, Claude 2 demonstrated an accuracy of 96.3% with an F1 score of 0.98. In instances where data were available, Claude 2 successfully extracted the pertinent information with a recall of 96.2% (151 out of 157 cases). Conversely, in situations where data were not reported in the reference standard, Claude 2 accurately reported the absence in 100% of the instances (3 out of 3 cases). Table 3 provides a 2×2 contingency table that illustrates the accuracy metrics. Table 4 presents the 16 data elements, their definitions, and the accuracy of extraction by Claude 2 for each data element. Appendix 3 presents the reference standard and the corresponding verbatim data extractions of Claude 2 for each data element in each of the 10 included studies.

**Table 3:**
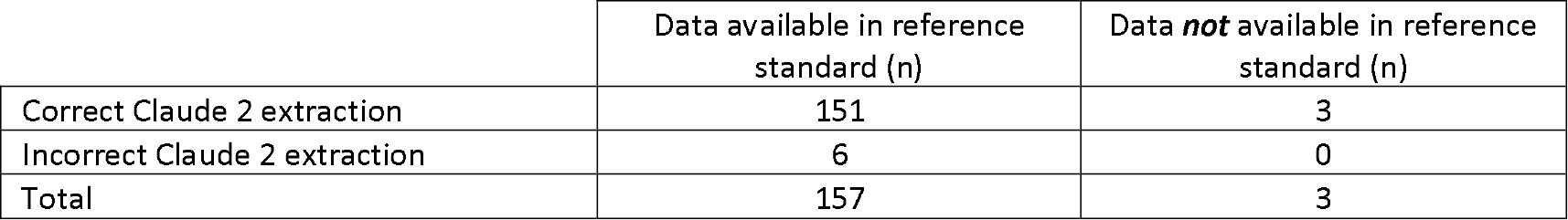
Contingency table of available data and performance of Claude 2 in data extraction.

**Table 4:** Elements for data extraction with definitions and accuracy of Claude 2.

| Data elements | Definitions | Accuracy |
| --- | --- | --- |
| **Study Identifiers** |  |  |
| First author, last name | The last name of the first author | 100% (10/10) |
| Trial registry number | The registry number of the study if one exists. Could be from any of several registries, including but not limited to clinicaltrials.gov | 90% (9/10) |
| Study name, acronym | The trial name and the acronym of the trial name if one exists | 100% (10/10) |
| Study funder | A text description of the study funder or sponsor, including multiple funders or sponsors if applicable. | 100% (10/10) |
| **Characteristics of Study Participants** |  |  |
| Mean age | The average age in years of study participants within each treatment group, reported to one decimal place | 90% (9/10) |
| Female participants | The total count and the corresponding percentage of female participants in each treatment group of the study, rounded to one decimal place | 100% (10/10) |
| Mean PASI score at baseline | The mean Psoriasis Area and Severity Index (PASI) score at baseline for participants in each treatment group | 100% (10/10) |
| Mean duration of disease | The average number of years that participants had plaque psoriasis, reported to one decimal place for each treatment group | 90% (9/10) |
| **Inclusion and Exclusion Criteria** |  |  |
| Inclusion criteria | A text description of the demographic, clinical, or other criteria used to select participants for entry into the study | 100% (10/10) |
| Exclusion criteria | A text description of the demographic, clinical, or other criteria used to exclude participants from the study | 90% (9/10) |
| **Numerical Data Related to Participant Flow** |  |  |
| N randomized | The total number of participants who were | 100% (10/10) |
|  | randomly assigned to any treatment group in the study |  |
| N randomized per group | The count of participants who were randomly assigned to each group of the study | 100% (10/10) |
| N analyzed | The count of participants who were analyzed in each group of the study | 100% (10/10) |
| **Intervention Characteristics** |  |  |
| Dose, route, frequency of intervention | The dose in milligram, the route of administration, and the frequency of the intervention for each treatment group | 100% (10/10) |
| **Outcome Data** |  |  |
| Primary outcome | Identification of the name and timing of the outcome designated as the primary outcome by the study authors | 100% (10/10) |
| Primary outcome, effect estimate | The effect estimate associated with the primary outcome for each treatment group; For dichotomous outcomes this could involve different forms of presentation of the effect estimate, e.g., counts and proportions, relative risks, odds ratios, hazard ratios, relative risk reductions, rate ratios, or absolute risk reductions. If reported, p-values should be presented as well. | 80% (8/10) |

Abbreviations: N = number of participants; PASI = Psoriasis Area and Severity Index

When we replicated data extractions to assess reliability, Claude 2 achieved 96.9% accuracy during replication 1, and 95.0% during replication 2.

### Reliability of data extractions

In two rounds of replications, we employed the final prompts on the same articles four weeks after the original data extraction to assess test-retest reliability. The proportions of errors remained low during both rounds of replication (replication 1: 3.1%; 5/160; replication 2: 5.0%; 8/160). It is worth noting that errors during replications largely occurred in different data items than those in the initial data extraction. The first replication shared just one data item with errors in common with the original extraction, while the second replication had two such common items.

### Types of errors

We categorized the six errors made by Claude 2 during the data extraction process using the categories outlined in Table 2. In four instances Claude 2 missed available data. For Papp et al.^21^, it reported the duration of disease by treatment group but failed to provide the overall duration of disease; for Lebwohl et al.^19^, it missed a p-value; for the study by Reich et al.^22^, Claude 2 failed to capture the study registration number and multiple inclusion and exclusion criteria. Additionally, it seemed to have generated extra participant exclusion criteria that we could not locate in either the study publication or the study registration (i.e., it hallucinated criteria, but we note this occurred after a revised prompt in response to some missing information in the response to our initial prompt).

In addition, we classified one instance as a major error. For the effect estimate of the primary outcome by Papp et al.^21^, Claude 2 extracted incorrect data for two groups: one set of data pertained to a different dosage group, while the other appeared to be fabricated. Finally, in one instance, we classified an error as a minor error. In the study by Thaçi et al,^26^ a standard deviation of the mean age of one treatment group was rounded incorrectly (should have been rounded to 14.0 not 13.9).

### Additional Observations

We observed several interesting aspects related to Claude 2’s performance. First, we noticed that in several cases Claude 2 was able to infer information not explicitly reported in the article. For example, our prompt related to the sex of participants specifically requested the percent female; in some studies Claude inferred this result based on the number of males enrolled and the total number of participants, which is something that our human extractors would have also had to have done to have reported the number and percent of females enrolled. Another example was the prompt related to number of participants analyzed. In many, but not all cases, Claude 2 correctly inferred the number of participants analyzed based on the number of participants randomized. In one case, the reference standard reported the median age or median duration of disease; in response to our prompts for the mean age and mean duration of disease, Claude 2 correctly told us that the mean was not reported but offered us the median, minimum, and maximum, which is the same inference our human extractors had made relative to the source PDF. Lastly, we observed that in comparison to our human extracted reference standard data, Claude 2 provided more complete and consistent responses, for example providing baseline characteristics by group and overall, consistently rounding to the same decimal place, standardized formatting of the extracted data, and including accent marks on author names where relevant. We did not consider these types of issues as errors in our human-extracted reference standard but rather areas where Claude-2 added value to the extraction process.

## Discussion

Our proof-of-concept study demonstrates the promising potential of leveraging LLMs, particularly Claude 2, for semi-automated data extraction for evidence syntheses. Claude 2 exhibited an impressive 96.3% accuracy in extracting data from publications of our selected studies. The test-retest reliability of Claude 2 across three rounds of data extraction was also high, although errors largely occurred in different data items during replications than in the initial data extraction. This indicates that the inherent stochasticity of LLMs has a minimal impact on the overall data extraction performance but plays a role for which items errors occur. Therefore, we were not able to determine from our sample whether any specific data items have a higher risk for errors. Across 3 rounds of automated data extraction with a total of 480 data elements, we encountered only seven major errors which could potentially affect estimates or conclusions of evidence syntheses. Twelve errors were minor or instances of missed data.

One noteworthy observation in our study was that Claude 2 required less prompt engineering than initially anticipated. It frequently provided responses that included the specific location in the article and verbatim text from which it extracted information, even though these details were not explicitly requested in the prompts. It accurately extracted data whether presented in text, figures, or tables.

A living review on the automation of data extraction has not detected any previous studies using LLMs for data extraction yet, so no direct comparison of the results of our proof-of-concept study with other studies assessing LLMs is possible.^9^ The high accuracy of Claude 2 in our study is difficult to compare with previous studies using other models because they often focused on data from abstracts alone, on the identification of sentences including relevant data, or on a few individual data items and not on a spectrum of data elements as used in evidence synthesis. A study using latent Dirichlet allocation along with logistic regression to extract inclusion and exclusion criteria, reported accuracies of 75% and 70%, respectively.^29^

Our study has several limitations. First, we used a convenience sample of 10 open-access RCTs from a single SR on drug treatment for a specific health condition, which does not fully represent the spectrum of study designs, interventions, and topic areas encountered in evidence syntheses. The applicability of our results to other study designs which may not have the structured reporting of RCTs, and to complex interventions, therefore, is unclear. Second, although we asked Claude 2 to extract different types of data, the tasks did not include continuous data, results from multiple study arms, and data that were not reported as primary outcomes. Third, our reference standard dataset contained only three instances where data we were interested in were not reported, constraining our evaluation of Claude 2’s performance in such scenarios and the risk of hallucinations. Consequently, the accuracy measures from our study should be interpreted with caution. The limited opportunity for false positive data extractions (i.e., hallucinations when no data are available) could potentially skew both, accuracy, and the F1-score. Across the three rounds of data extraction (original and replications), however, we encountered only two instances of fabricated data (i.e., hallucinations).

Future research needs to focus on the development of use cases for Claude 2 in evidence synthesis so that investigators can get a more comprehensive understanding of its capabilities and limitations. Such use cases need to assess Claude 2’s performance in extracting data from non-randomized study designs, complex interventions, continuous outcomes, and data that are not reported as primary outcomes. They should also assess whether Claude 2 can perform simple calculations that are common during the data extraction process (e.g., the calculation of counts from proportions and vice versa). Ideally, such use cases would include more instances of missing data (i.e., relevant data that are not reported in study publications) than our study did. Additionally, it is crucial to investigate the time and resource efficiencies gained from utilizing LLMs for data extraction compared to traditional human-led data extraction. Such a prospective comparison should also consider factors such as prompt engineering, data curation, as well as the pilot testing and double-checking of results on the human side. Moreover, we suggest exploring the stability of LLMs over time, as models like Claude 2 and others are continually evolving. Investigating whether the same prompts yield consistent results over an extended period is essential for assessing the reliability of these tools for evidence syntheses. Lastly, future research should compare the performance of Claude 2 with other LLM models, such as GPT-4 for different types of data and study designs.

The potential implications of our study for evidence syntheses are significant. Leveraging LLMs like Claude 2 not only promises to enhance efficiency in data extraction but also to improve the overall accuracy of this critical process. An unexpected finding from our study were the number of errors present in our human-extracted reference standard. By reducing human error, these models can contribute to more robust evidence synthesis products. A major strength of Claude 2 and other LLMs is their exceptional usability. The web-browser interface of LLMs make them user-friendly for researchers without a technical background. In contrast, prior studies on automated data extraction show that nearly 90% of models necessitate pre-processing the text data before employing algorithms for data extraction.^9^ Furthermore, LLMs can perform data extraction on any topic without requiring a labeled training set. Claude 2 enables easy PDF uploads in the context window without necessitating a format conversion or text parsing.

Nonetheless, the integration of LLMs in the data extraction process of evidence synthesis should for now, only be done in the form of semi-automation. When an LLM extracts data, it remains essential for a human investigator to validate the data, akin to the role of a second investigator in traditional human-led data extraction. However, it is possible that over time these LLMs could be used to check their own work in a way that even outperforms humans.

Not surprisingly, our study further underscores the fallibility of human data extractors, whose previously extracted data emerged as an imperfect reference standard in our study. Utilizing LLMs for data extraction offers the opportunity to redirect human effort from monotonous, error-prone work to tasks that require human judgment such as the understanding the context of the evidence, synthesis and interpretation of evidence, and stakeholder engagement.

In conclusion, the synergy between human expertise and LLMs has the potential to revolutionize data extraction and ultimately improve the quality and efficiency of evidence synthesis products.

## Ethical Considerations

The study is solely aimed at advancing knowledge and understanding in the field of AI-based data extraction, without involving any human subjects or sensitive data. Therefore, ethical considerations and associated procedures are not applicable for this research.

## Acknowledgements

Thank you to Colleen Ovelman for her support during the project’s initial stages, Petra Wellemsen of Danube University, Krems for manuscript formatting, and the Innovation Team at RTI International for their project support.

## Data Availability

Data supporting the findings of this study are available from the corresponding author upon reasonable request.

## Author contributions

Conceptualization: GG, KC, MV, LK; Funding acquisition: GG, KC; Project administration: GB; Investigation: KC, MV, LK, NE, RH, BNS, SK, IT, AK, GB, GG; Data curation: SK, AK; Writing original draft: GG, RC; Review and revisions of draft: KC, MV, LK, NE, RH, BNS, SK, IT, AK, GB.

