## Appendix for "Data Extraction for Evidence Synthesis Using a Large Language Model: A Proof-of-Concept Study"

### Appendix 1

**Data elements and respective prompts used for Claude 2 to extract data from journal publications**

| **Data elements** | **Prompt** |
| --- | --- |
| First author's last name | State the last name of the first author, styled as a proper noun with first letter capitalized. |
| Trial registry number | State the registry number of the study, if one exists. Could be from any of a number of registries, including but not limited to clinicaltrials.gov |
| Study name and acronym | State the trial name and the acronym of the trial name, if one exists. |
| Study funder | State the study funder or sponsor, including multiple funders or sponsors if applicable. |
| Mean age | State the average age in years with standard deviation, reported to two decimal place, of study participants within each treatment group. |
| Female participants | State the total count and the corresponding percentage, rounded to one decimal place, of female participants in each treatment group of the study. |
| Mean PASI score at baseline | State the mean Psoriasis Area and Severity Index (PASI) score with standard deviation at baseline for participants in each treatment group. |
| Mean duration of disease | State the average number of years with standard deviation reported to one decimal place that participants had plaque psoriasis for each treatment group. |
| Inclusion criteria | State only the study inclusion criteria. |
| Exclusion criteria | State the study exclusion criteria. |
| N_randomized | State the total number of participants who were randomly assigned to any treatment group in the study. |
| N_randomized_per_group | State the count of participants who were randomly assigned to each group of the study. |
| N_analyzed | State the count of participants who were analyzed in each group of the study. |
| Dose_route_frequency | State the dose in milligram, the route of administration, and the frequency of the intervention for each treatment. |
| Primary outcome_name | State the name and/or timing of the outcome designated as the primary outcome by the study authors. |
| Primary outcome_estimate | State the effect estimate associated with the primary outcome for each treatment group. For dichotomous outcomes this could involve different forms of presentation of the effect estimate, e.g., counts and proportions, relative risks, odds ratios, hazard ratios, relative risk reductions, rate ratios, or absolute risk reductions. If reported, p-values should be presented as well. |

### Appendix 2

**Errors in human-extracted reference standard that were corrected during the proof-of-concept study (N=21)**

| **Study** | **Data extraction in original reference standard** | **Data extraction in corrected reference standard** | **Type of error** |
| --- | --- | --- | --- |
| Bagel et al., 2018 (CLARITY)^1^ | Three issues:  *N (%) Female Participants*  Ustekinumab: 176 (***31.8***)  *IGA 0/1 response achieved* Ustekinumab: ***264*** (55.4)  *Exclusion criteria*  NR | Three issues:  N (%) Female Participants  Ustekinumab: 176 (***31.9***)  *IGA 0/1 response achieved*  Ustekinumab: ***306*** (55.4)  *Exclusion criteria*  Forms of psoriasis other than plaque psoriasis, drug-induced psoriasis, ongoing use of prohibited treatments, previous exposure to secukinumab or any other biologic drug directly targeting IL17A or IL-17RA, or ustekinumab, or any therapies targeting IL-12 or IL-23 5. | Three issues:  Rounding error in the proportion  Frequency incorrectly transcribed  Data omitted |
| Blauvelt et al., 2020^2^ | Two issues:  *N analyzed*  Ixekizumab: 520  Guselkumab: 507  *Dosage*  Ixekizumab: ***80 mg, subcutaneous, every 2 weeks*** through week 12, then every 4 weeks; Ustekinumab: 45 or 90 mg (if patient weight more than 100 kg), subcutaneous, weeks 0, 4, and 16 | Two issues:  *N analyzed*  -Ixekizumab: 520 (efficacy), ***519 (safety)***  Guselkumab: 507 (efficacy), ***506 (safety)***  *Dosage*  Ixekizumab:  - Dose: ***160 mg at week 0, then 80 mg***  - Route: Subcutaneous injection  - Frequency: Every 2 weeks through week 12, then every 4 weeks | Two issues:  Only abstracted the N analyzed for efficacy.  Loading dose for one of the drugs missing. |
| Glatt et al., 2016^3^ | Four issues:  *Mean age (SD)* Placebo: 38.2 (13.3)  ***Bimekizumab: 39.5*** *(10.4)*  *N (%) Female*  Placebo: 1 (***7.3***)  Bimekizumab: 9 (23.1)  *Inclusion criteria*  Adults ages 18 to 70 with plaque-type psoriasis for at least 6 months involving ***more than 5% of BSA*** (excluding the scalp) and at least 2 psoriatic lesions with at least 1 plaque in a suitable biopsy site.  *N analyzed*  Bimekizumab 8 mg: ***3***  Bimekizumab 40 mg: 4 Bimekizumab 160 mg: 6 Bimekizumab 480 mg: ***5*** Bimekizumab 640 mg: 6  Placebo: 13 | Four issues:  *Mean age (SD)*  ***Overall: 39.5*** (10.4)  Placebo: 38.2 (13.3)  Bimekizumab 8 mg: 34.7 (9.7)  Bimekizumab 40 mg: 44.8 (10.5)  Bimekizumab 160 mg: 43.5 (8.4)  Bimekizumab 480 mg: 39.5 (9.1)  Bimekizumab 640 mg: 38.1 (7.2)  *N (%) Female*  Placebo: 1 (***7.7***)  Bimekizumab: 9 (23.1)  *Inclusion criteria*  Adults aged 18 to 70 years with plaque-type psoriasis for at least 6 months ***with BSA of 5% or less*** excluding the scalp, at least 2 psoriatic lesions with at least 1 plaque in a suitable biopsy site.  *N analyzed*  Bimekizumab 8 mg: ***4*** Bimekizumab 40 mg: 4 Bimekizumab 160 mg: 6 Bimekizumab 480 mg: ***6*** Bimekizumab 640 mg: 6 Placebo: 13 | Four issues:  Humans incorrectly attributed the data for all study participants to bimekizumab users only.  Rounding error in the proportion  Transcribed part of criteria incorrectly.  Humans incorrectly abstracted the number completed rather than the number analyzed for 2 of the dosage groups. |
| Lebwohl et al. 2018^4^ | *N (%) Female*  Placebo: 23 (60.4) | *N (%) Female*  Placebo: 23 (40.4) | Proportion was based on number of males instead of females. |
| Reich et al, 2017 (LIBERATE)^5^ | Four issues:  *Dosage*  Etanercept 50 mg subcutaneous ***twice*** weekly  *Mean (SD) PASI Score*  NR  *Mean (SD) duration of disease* NR  *Exclusion criteria*  NR | Four issues:  *Dosage*  Etanercept 50 mg ***once*** weekly  *Mean (SD) PASI Score*  Placebo: 19.4 (6.8)  Apremilast: 19.3 (7.0)  Etanercept: 20.3 (7.9)  *Mean (SD) duration of disease*  Placebo: 16.6 (12.1)  Apremilast: 19.7 (12.7)  Etanercept: 18.1 (11.7)  *Exclusion criteria*  Prior exposure to a biologic therapy, prior failure of more than 3 systemic agents for treatment of psoriasis, history of known demyelinating diseases such as multiple sclerosis or optic neuritis, history of or concurrent congestive heart failure, including medically controlled, asymptomatic congestive heart failure, other clinically significant or major uncontrolled disease, serious infection, latent, active or history of incompletely treated tuberculosis | Four issues:  Wrong frequency of dosing  Data omitted  Data omitted  Data omitted |
| Reich et al., 2017^6^ | *Dosage*  Guselkumab: 100 mg, subcutaneous, weeks 0, 4, 12 | *Dosage*  Guselkumab: 100 mg, subcutaneous, weeks 0, 4, 12, ***and 20*** | Omitted the final dosage |
| Reich et al., 2017 (IXORA-S)^7^ | Two issues:  *Mean duration of disease*  Ixekizumab: 18.0 (***1.1***)  *Exclusion criteria*  NR | Two issues:  *Mean duration of disease*  Ixekizumab: 18.0 (***11.1***)  *Exclusion criteria*  Predominant presence of nonplaque psoriasis, a contraindication for ustekinumab, or prior treatment with ustekinumab, ixekizumab or any other IL-17 or IL-12/23  antagonists. | Two issues:  Incorrectly transcribed numeric value for SD  Data omitted |
| Thaci et al. (2015) ^8^ | Four issues:  *Mean (SD) PASI Score*  NR  *Mean (SD) duration of disease* NR  *Exclusion criteria*  NR  *Dosage*  Secukinumab 300 mg SC at ***weeks 0, 1, 2, and 3*** then every 4 weeks | Four issues:  *Mean (SD) PASI Score*  Secukinumab: 21.7 (8.50) Ustekinumab: 21.5 (8.07)  *Mean (SD) duration of disease*  Secukinumab: 19.6 (12.90) Ustekinumab: 16.1 (11.24)  *Exclusion criteria*  Prior exposure to any biologics directly targeting IL-17A/IL-17 receptor A or IL-12/IL-23.  *Dosage*  Secukinumab 300 mg SC ***at weeks 0, 1, 2, 3 and 4 then*** every 4 weeks | Four issues:  Data omitted  Data omitted  Data omitted  Error in dosing interval |

### Appendix 3

**Comparison of data of reference standard and data extracted by Claude 2 using final prompts for each data element by journal publication**

**Bagel et al. 2018^1^**

| **Variable** | **Reference standard** | **Claude 2^9^** |
| --- | --- | --- |
| First author's last name | Bagel | The last name of the first author is Bagel. |
| Trial registry number | NCT02826603 | The clinicaltrials.gov identifier for this study is NCT02826603. This is stated in the Methods section:  "CLARITY (NCT02826603) is a multicenter, randomized, double-blinded, active-controlled, parallel-group, phase 3b trial." |
| Study name and acronym | CLARITY | The trial name is "Secukinumab is Superior to Ustekinumab in Clearing Skin in Patients with Moderate to Severe Plaque Psoriasis (16-Week CLARITY Results)".   The acronym for the trial name is CLARITY. |
| Study funder | Novartis Pharma AG | The study was funded by Novartis Pharma AG, Basel, Switzerland. This is stated in the Funding section:  "Funding. This investigation was sponsored by Novartis Pharma AG, Basel, Switzerland. Novartis Pharma AG also funded the journal article processing charges."  So the sole study funder/sponsor was Novartis Pharma AG. |
| Mean age | Secukinumab: 45.4 (14.1); Ustekinumab: 45.3 (14.2) | According to Table 1 in the paper, the average age in years with standard deviation reported to one decimal place was:  Secukinumab group: 45.4 ± 14.1 Ustekinumab group: 45.3 ± 14.2 |
| Female participants | Secukinumab: 194 (35.3); Ustekinumab: 176 (31.8) | The total number and percentage of female participants in each treatment group is not reported separately. However, the overall number of male participants is provided in Table 1:  - Secukinumab group: 356 males (64.7% of 550)  - Ustekinumab group: 376 males (68.1% of 552)  Since the total number of participants in each group is provided, I can calculate the number and percentage of females as:  - Secukinumab group: 194 females (35.3% of 550) - Ustekinumab group: 176 females (31.9% of 552)  To one decimal place, the percentage of female participants in each group was:  - Secukinumab: 35.3%  - Ustekinumab: 31.9% |
| Mean PASI score at baseline | Secukinumab: 20.8 (9.0); Ustekinumab: 21.3 (9.2) | Secukinumab group: 45.4 ± 14.1 |
| Mean duration of disease | Secukinumab: 16.8 (11.9); Ustekinumab: 17.3 (13.3) | According to Table 1 in the paper, the average number of years with standard deviation reported to one decimal place that participants had plaque psoriasis was:  Secukinumab group: 16.8 ± 11.9 years Ustekinumab group: 17.3 ± 13.3 years |
| Inclusion criteria | Adults aged 18 and older with moderate-to-severe chronic plaque psoriasis defined by PASI ≥ 12, static 5-point IGA 2011 modified version score ≥ 3, and BSA involvement ≥ 10%, inadequately controlled by topical treatments, phototherapy, and/or previous systemic therapy. | The key inclusion criteria for the study are not fully listed in the abstract, but are described in the Methods section:  "Patients (≥18 years) with moderate to severe chronic plaque psoriasis (PASI ≥12, static 5-point Investigator’s Global Assessment 2011 modified version [IGA mod 2011] score ≥3, and body surface area [BSA] involvement ≥10%) and who were inadequately controlled by topical treatments, phototherapy, and/or previous systemic therapy were eligible."  So the key inclusion criteria were: - Adults aged ≥18 years - Moderate to severe chronic plaque psoriasis - PASI score ≥12  - IGA modified 2011 score ≥3 - BSA involvement ≥10% - Inadequate control with prior therapies including topicals, phototherapy, and/or systemic treatments |
| Exclusion criteria | Forms of psoriasis other than plaque psoriasis, drug-induced psoriasis, ongoing use of prohibited treatments, previous exposure to secukinumab or any other biologic drug directly targeting IL17A or IL-17RA, or ustekinumab, or any therapies targeting IL-12 or IL-23 5, use of any other investigational drugs within 5 half-lives of the investigational treatment before study drug initiation, or pregnant or nursing women  from the supplementary file. not in the evidence table. | Based on the provided Supplementary Table 1, the key exclusion criteria for the study are:  1. Forms of psoriasis other than plaque psoriasis  2. Drug-induced psoriasis  3. Ongoing use of prohibited treatments  4. Previous exposure to secukinumab, other biologics directly targeting IL-17A or IL-17RA, ustekinumab, or any therapies targeting IL-12 or IL-23  5. Use of any other investigational drugs within 5 half-lives before study drug initiation  6. Pregnant or nursing (lactating) women |
| N_randomized | 1102 | The total number of participants randomized in the study is provided in the Methods section:  "A total of 1102 patients, of whom almost two-thirds were US patients (64.2%), were randomized to either secukinumab 300 mg (n = 550) or ustekinumab 45/90 mg (n = 552)."  So the total number of participants randomly assigned to any treatment group was 1102. |
| N_randomized_per_group | Secukinumab: 550; Ustekinumab: 552 | The number of participants randomly assigned to each treatment group is provided in the Methods section:  "A total of 1102 patients, of whom almost two-thirds were US patients (64.2%), were randomized to either secukinumab 300 mg (n = 550) or ustekinumab 45/90 mg (n = 552)."  So the number of participants randomly assigned to each group was:  - Secukinumab 300 mg: 550 - Ustekinumab 45/90 mg: 552 |
| N_analyzed | Secukinumab: 550; Ustekinumab: 552  (imputation used; number completed is different) | The number of participants analyzed in each treatment group is not explicitly stated in the abstract. However, the Results section indicates that the analysis was based on the randomized populations:  "Secukinumab was superior to ustekinumab for the proportion of patients that achieved PASI 90 responses at Week 12 (66.5% vs. 47.9%; p < 0.0001) and for the proportion of patients that achieved IGA mod 2011 0/1 responses at Week 12 (72.3% vs. 55.4%; p < 0.0001)"  Since the analysis is based on the randomized populations, I can infer that the number of participants analyzed in each group was the same as the number randomized, which was:  - Secukinumab 300 mg: 550  - Ustekinumab 45/90 mg: 552 |
| Dose_route_frequency | Secukinumab: 300 mg, subcutaneous, weeks 0, 1, 2, and 3 and then every 4 weeks from weeks 4 to 48; Ustekinumab: 45 mg for patient weighing 100 kg or less, 90 mg for patient weighing more than 100 kg, subcutaneous, weeks 0, 4, 16, 28, and 40 | The dose, route of administration, and frequency for each treatment is provided in the Methods section:  "Eligible patients were randomized 1:1 to receive either subcutaneous secukinumab 300 mg at Baseline, Weeks 1, 2, and 3, and then every 4 weeks from Weeks 4 to 48, or subcutaneous ustekinumab (45 mg for patient weighing ≤100 kg or 90 mg for patient weighing >100 kg) at Baseline, Week 4, and then every 12 weeks."  So the details for each treatment are:  Secukinumab: - Dose: 300 mg  - Route: Subcutaneous - Frequency: Baseline, Weeks 1, 2, and 3, then every 4 weeks  Ustekinumab:  - Dose: 45 mg if ≤100 kg, 90 mg if >100 kg - Route: Subcutaneous - Frequency: Baseline, Week 4, then every 12 weeks |
| Primary outcome_name | PASI 90 response; IGA mod 2011 0/1 (clear or almost clear skin) | The primary outcomes of the study are stated in the Methods section:  "The co-primary objectives of the study were to demonstrate the superiority of secukinumab compared to ustekinumab with respect to PASI 90 response and IGA mod 2011 0/1 (clear or almost clear skin) at Week 12."  So the primary outcomes designated by the study authors were:  - PASI 90 response at Week 12 - IGA mod 2011 0/1 response at Week 12 |
| Primary outcome_estimate | PASI 90: Secukinumab: 366 (66.5); Ustekinumab: 264 (47.9); p<0.0001; IGA mod 2011 0/1 response achieved: Secukinumab: 398 (72.3); Ustekinumab: 264 (55.4); p<0.0001  co-primary outcomes | For the co-primary endpoints at week 12:  PASI 90 response: - Secukinumab group: 366/550 (66.5%)  - Ustekinumab group: 264/552 (47.9%) - Odds ratio was 2.16 (95% CI 1.71, 2.72), p<0.0001 in favor of secukinumab  IGA 0/1 response:  - Secukinumab group: 398/550 (72.3%) - Ustekinumab group: 306/552 (55.4%)  - Odds ratio was 2.13 (95% CI 1.65, 2.75), p<0.0001 in favor of secukinumab  So secukinumab showed significantly higher response rates compared to ustekinumab on both co-primary endpoints. The odds of achieving PASI 90 and IGA 0/1 responses were approximately 2-fold higher with secukinumab compared to ustekinumab. |

**Blauvelt et al. 2020^2^**

| **Variable** | **Reference standard** | **Claude 2^9^** |
| --- | --- | --- |
| First author's last name | Blauvelt | The last name of the first author is Blauvelt. |
| Trial registry number | NCT03573323 | The clinical trial registry number for this study is NCT03573323 on ClinicalTrials.gov. |
| Study name and acronym | IXORA-R | The trial name is IXORA-R. IXORA-R stands for "Ixekizumab Compared to Guselkumab in Patients With Moderate to Severe Plaque Psoriasis". |
| Study funder | Eli Lilly and Company | The study was funded by Eli Lilly and Company. |
| Mean age | Ixekizumab: 49.0 (13.9); Guselkumab: 49.0 (14.9) | According to Table 1 in the paper, the average age and standard deviation for each treatment group was:  - Ixekizumab group: 49.0 ± 13.9 years - Guselkumab group: 49.0 ± 14.9 years  Reporting the mean and standard deviation to one decimal place:  - Ixekizumab group: 49.0 ± 13.9 years - Guselkumab group: 49.0 ± 14.9 years |
| Female participants | Ixekizumab: 182 (35); Guselkumab: 193 (38) | According to Table 1:  - In the ixekizumab group, there were 182 female participants out of 520 total, which is 35.0% female. - In the guselkumab group, there were 193 female participants out of 507 total, which is 38.1% female.  Rounded to one decimal place: - Ixekizumab group: 35.0% female  - Guselkumab group: 38.1% female |
| Mean PASI score at baseline | Ixekizumab: 19.5 (7.9); Guselkumab: 19.3 (7.1) | According to Table 1, the mean PASI score and standard deviation at baseline for each treatment group was:  - Ixekizumab group: mean PASI = 19.5, SD = 7.9 - Guselkumab group: mean PASI = 19.3, SD = 7.1 |
| Mean duration of disease | Ixekizumab: 17.5 (13.8); Guselkumab: 16.3 (13.8) | According to Table 1, the average number of years since diagnosis of plaque psoriasis and standard deviation for each treatment group was:  - Ixekizumab group: 17.5 ± 13.8 years - Guselkumab group: 16.3 ± 13.8 years  Reporting the mean and standard deviation to one decimal place:  - Ixekizumab group: 17.5 ± 13.8 years - Guselkumab group: 16.3 ± 13.8 years |
| Inclusion criteria | Adults aged 18 and older with moderate-to-severe plaque psoriasis for a minimum of 6 months’ duration defined by PASI ≥ 12, PGA ≥ 3 on a 5-point scale, and BSA involvement ≥ 10% and candidate for systemic therapy. | According to the Patients and Methods section, the inclusion criteria for the study were:  - Adults aged ≥18 years  - Diagnosis of chronic plaque psoriasis for at least 6 months before baseline, as determined by the investigator - Candidates for phototherapy and/or systemic therapy - Static Physician's Global Assessment (sPGA) score of ≥3  - PASI score ≥12 - ≥10% body surface area involvement |
| Exclusion criteria | Nonplaque psoriasis, history of drug-induced psoriasis, used a tanning booth 4 weeks before baseline, used any biological agent within specified periods prior to baseline, used any IL-23p19 antagonists, or had any condition as addressed in the local labeling for guselkumab. Prior use of an IL-17 antagonist permitted if the patient had not failed to respond to the therapy. | According to the Patients and Methods section, the exclusion criteria for the study were:  - Predominant pustular, erythrodermic and/or guttate forms of psoriasis  - History of drug-induced psoriasis or a clinically significant flare of psoriasis during the 12 weeks before baseline - Use of tanning booths 4 weeks before baseline - Use of any biological agent within specified periods prior to baseline - Any use of IL-23p19 antagonists - Any condition or contraindication addressed in the local labelling for guselkumab - Prior use of an IL-17 antagonist other than ixekizumab if the patient failed to respond to the therapy |
| N_randomized | 1027 | According to the Results section, a total of 1027 patients were randomized to receive treatment - 507 to guselkumab and 520 to ixekizumab. |
| N_randomized_per_group | Ixekizumab: 520; Guselkumab: 507 | According to the Results section:  - 507 participants were randomly assigned to the guselkumab group - 520 participants were randomly assigned to the ixekizumab group |
| N_analyzed | Ixekizumab: 520; Guselkumab: 507 | The efficacy analyses included all randomized patients according to the treatment they were assigned (intent-to-treat population). The safety data included all randomized patients who received at least one dose of the trial drug (safety population).  The number of patients in each group were:  - Ixekizumab group: 520 (efficacy), 519 (safety)  - Guselkumab group: 507 (efficacy), 506 (safety)  So 520 participants in the ixekizumab group and 507 participants in the guselkumab group were analyzed for efficacy. 519 participants in the ixekizumab group and 506 participants in the guselkumab group were analyzed for safety. |
| Dose_route_frequency | Ixekizumab: 80 mg, subcutaneous, every 2 weeks then every 4 weeks; Guselkumab: 100 mg, subcutaneous, weeks 0, 4, 12, and 20 | According to the Procedures section:  Ixekizumab: - Dose: 160 mg starting dose, then 80 mg  - Route: Subcutaneous injection - Frequency: 160 mg starting dose at week 0 (two 80 mg injections), then 80 mg every 2 weeks from weeks 2 to 12  Guselkumab:  - Dose: 100 mg - Route: Subcutaneous injection  - Frequency: 100 mg injections at weeks 0, 4, and 12 |
| Primary outcome_name | PASI 100 | According to the Outcomes section, the primary efficacy endpoint was the percentage of patients reaching 100% improvement from baseline in PASI score (PASI 100) at week 12.  So the primary outcome was PASI 100 response at 12 weeks. |
| Primary outcome_estimate | Ixekizumab: 215 (41); Guselkumab: 126 (25); OR: 2.14, 95% CI: 1.63 to 2.81, p< 0.001 | For the primary outcome of PASI 100 at 12 weeks:  - Ixekizumab group: 41% (215/520 patients)  - Guselkumab group: 25% (126/507 patients)  The odds ratio for ixekizumab vs. guselkumab was 2.14 (95% CI 1.63-2.81), p<0.001.  So ixekizumab showed a significantly higher proportion of patients achieving PASI 100 compared to guselkumab at 12 weeks. |

**Glatt et al. 2016^3^**

| **Variable** | **Reference standard** | **Claude 2^9^** |
| --- | --- | --- |
| First author's last name | Glatt | The last name of the first author is Glatt. |
| Trial registry number | NCT02529956 | The clinical trial registry number for this study, as stated in the paper, is NCT02529956 on clinicaltrials.gov. |
| Study name and acronym |  | The trial name is not explicitly stated in the paper. However, based on the title and content of the paper, the trial name appears to be "First-in-human randomized study of bimekizumab, a humanized monoclonal antibody and selective dual inhibitor of IL-17A and IL-17F, in mild psoriasis". There is no acronym mentioned for the trial name. |
| Study funder | UCB Pharma | The study was funded by UCB Pharma, as stated in the "Competing Interests" section:  "The study was funded by UCB Pharma. Additional statistical support was provided by Emma Jones of Veramed Ltd and Ros Walley of UCB."  So the primary study funder/sponsor was UCB Pharma. |
| Mean age | Placebo: 38.2 (13.3); Bimekizumab: 39.5 (10.4) | The average age of study participants in each treatment group is not reported in the paper. The only age information provided is the overall mean age across all participants, which was 39.53 years. The paper does not report mean or average age broken down by each treatment group. |
| Female participants | Placebo: 1 (7.3); Bimekizumab: 9 (23.1) | Based on the total population:  Placebo: 1 female, 7.7%  Total: 9 females, 23.1%  Based solely on the male population:  Placebo: 1 female, 7.7% Total: 9 females, 30.0% |
| Mean PASI score at baseline | Placebo: 3.0 (1.8 to 6.1); Bimekizumab: 3.5 (0.8 to 6.7) | The paper provides the following PASI information at baseline:  Placebo:  - Median PASI = 3.00 - Min PASI = 1.8 - Max PASI = 6.1  Total: - Median PASI = 3.50  - Min PASI = 0.8 - Max PASI = 6.7 |
| Mean duration of disease | Not reported | The paper does not report the average number of years that participants had plaque psoriasis for each treatment group. The only information provided about duration of psoriasis is in the inclusion criteria, which states:  "Subjects with a confirmed diagnosis of mild-to-moderate plaque-type psoriasis for ≥6 months involving ≤5% of body surface area (excluding the scalp)."  So the minimum duration of psoriasis for inclusion in the study overall was 6 months, but the average duration for each treatment group is not reported. |
| Inclusion criteria | Adults aged 18 to 70 years with plaque-type psoriasis for at least 6 months with BSA of 5% or less excluding the scalp, at least 2 psoriatic lesions with at least 1 plaque in a suitable biopsy site. | The key inclusion criteria for the study, as stated in the Methods section, were:  - Male or female (age ≥18 to ≤70 years) - Subjects with a confirmed diagnosis of mild-to-moderate plaque-type psoriasis for ≥6 months involving ≤5% of body surface area (excluding the scalp)  - At least two psoriatic lesions with at least one plaque in suitable biopsy sites |
| Exclusion criteria | Use of systemic nonbiological psoriasis therapy or phototherapy within 4 weeks prior to screening or treatment with biological agents within 12 months prior to screening. | The key exclusion criteria for the study, as stated in the Methods section, were:  - Use of systemic nonbiological psoriasis therapy (methotrexate, steroids or cyclophosphamide) or psoralen plus UVA/UVB phototherapy within 4 weeks prior to screening - Treatment with biological agents ≤12 months before the study |
| N_randomized | 39 | The total number of participants who were randomly assigned to a treatment group in the study was 39, as stated in the Results section:  "Thirty-nine (comprising the full analysis set [FAS] and PK -per-protocol set) were randomized to treatment (bimekizumab, n = 26; placebo, n = 13)."  So the total number of participants randomly assigned to any treatment group was 39. |
| N_randomized_per_group | Bimekizumab 8 mg: 4; Bimekizumab 40 mg: 4; Bimekizumab 160 mg: 6; Bimekizumab 480 mg: 6; Bimekizumab 640 mg: 6; Placebo: 13 | Based on the information provided in the Results section, the number of participants randomly assigned to each group was:  - Placebo: 13 - Bimekizumab 8 mg: 4  - Bimekizumab 40 mg: 4 - Bimekizumab 160 mg: 6 - Bimekizumab 480 mg: 6  - Bimekizumab 640 mg: 6  So the count of participants randomly assigned to each group was:  Placebo: 13 Bimekizumab 8 mg: 4 Bimekizumab 40 mg: 4  Bimekizumab 160 mg: 6 Bimekizumab 480 mg: 6 Bimekizumab 640 mg: 6 |
| N_analyzed | Bimekizumab 8 mg: 3; Bimekizumab 40 mg: 4; Bimekizumab 160 mg: 6; Bimekizumab 480 mg: 5; Bimekizumab 640 mg: 6; Placebo: 13 | The number of participants analyzed in each group is not explicitly stated in the paper. However, based on the information provided, the full analysis set (FAS) comprised all 39 randomized subjects. Therefore, the number of participants analyzed in each group can be assumed to be the same as the number randomized to each group, which was:  - Placebo: 13 - Bimekizumab 8 mg: 4  - Bimekizumab 40 mg: 4 - Bimekizumab 160 mg: 6 - Bimekizumab 480 mg: 6 - Bimekizumab 640 mg: 6  So the count of participants analyzed in each group appears to be the same as those randomized:  Placebo: 13  Bimekizumab 8 mg: 4 Bimekizumab 40 mg: 4 Bimekizumab 160 mg: 6  Bimekizumab 480 mg: 6 Bimekizumab 640 mg: 6 |
| Dose_route_frequency | Bimekizumab: 8 mg, IV, one time; 40 mg, IV, one time; 160 mg, IV, one time; 480 mg, IV, one time; 640 mg, IV, one time | Based on the information provided in the Methods section:  - Placebo group: Single intravenous infusion of placebo  - Bimekizumab groups:  - 8 mg group: Single intravenous infusion of 8 mg bimekizumab  - 40 mg group: Single intravenous infusion of 40 mg bimekizumab   - 160 mg group: Single intravenous infusion of 160 mg bimekizumab  - 480 mg group: Single intravenous infusion of 480 mg bimekizumab  - 640 mg group: Single intravenous infusion of 640 mg bimekizumab  So in summary, the dose, route and frequency for each treatment was:  - Placebo: Single intravenous infusion  - Bimekizumab 8 mg: Single intravenous infusion  - Bimekizumab 40 mg: Single intravenous infusion - Bimekizumab 160 mg: Single intravenous infusion - Bimekizumab 480 mg: Single intravenous infusion - Bimekizumab 640 mg: Single intravenous infusion |
| Primary outcome_name | Safety   co-primary outcomes | The primary outcome of the study was to evaluate the safety of single-dose bimekizumab, as stated in the Objectives section:  "The primary objective was to evaluate the safety of single-dose bimekizumab."  The safety assessments were done at various timepoints during and after the single infusion, as described in the Methods section under Safety outcomes.  So the primary outcome was safety of single-dose bimekizumab, assessed at multiple timepoints during and after the infusion. |
| Primary outcome_estimate | TEAEs: Bimekizumab: 22 (84.6); placebo: 10 (76.9);  Treatment-related TEAEs: Bimekizumab: 12 (46.2); placebo: 4 (30.8);  SAEs: Bimekizumab: 1 (3.8); placebo: 0 (0);   Injection-site reactions: Bimekizumab: 0; placebo: 0  (Did not include calculated RRs; evidence table indicates NR for injection site reactions - publication notes none reported.) | Since this was a Phase 1 first-in-human study, the primary outcome was to evaluate the safety and tolerability of bimekizumab. There was no single predefined efficacy outcome.  Here is a summary of the key safety findings:   Adverse events:  Bimekizumab: 22/26 (84.6%) subjects  Placebo: 10/13 (76.9%) subjects  Serious adverse events:  Bimekizumab: 1/26 (3.8%) subjects  Placebo: 0/13 (0%) subjects  Discontinuations due to adverse events:  Bimekizumab: 0/26 (0%)  Placebo: 0/13 (0%)  Severe adverse events:  Bimekizumab: 0/26 (0%)  Placebo: 0/13 (0%)  No quantitative effect estimates like relative risks or odds ratios were reported for the safety outcomes.  Overall, bimekizumab was considered well tolerated across the doses studied. The study did not identify any significant safety signals or concerns. |

**Lebwohl et al. 2018^4^**

| **Variable** | **Reference standard** | **Claude 2^9^** |
| --- | --- | --- |
| First author's last name | Lebwohl | The last name of the first author is Lebwohl. |
| Trial registry number | NCT02346240 | According to the manuscript, this study is registered at ClinicalTrials.gov with identifier NCT02346240. |
| Study name and acronym | CIMPACT | The trial name is CIMPACT (CIMPASI). No acronym is explicitly stated for the trial name in the manuscript. |
| Study funder | Dermira Inc and UCB Inc | The study was funded by Dermira Inc and UCB Inc. According to the manuscript, UCB is the regulatory sponsor of certolizumab pegol in psoriasis. |
| Mean age | Certolizumab pegol 200 mg: 46.7 (13.5); Certolizumab pegol 400 mg: 45.4 (12.4); Etanercept: 44.6 (14.1); Placebo: 46.5 (12.5) | Based on the information provided in Table I of the manuscript:  - Placebo group: Average age = 46.5 years - Etanercept group: Average age = 44.6 years  - CZP 200 mg Q2W group: Average age = 46.7 years - CZP 400 mg Q2W group: Average age = 45.4 years |
| Female participants | Certolizumab pegol 200 mg: 52 (31.5); Certolizumab pegol 400 mg: 60 (35.9); Etanercept: 43 (25.3); Placebo: 23 (60.4) | The manuscript does not explicitly state the number or percentage of female participants in each treatment group. However, Table I provides the number of male participants in each group:  - Placebo: 34 males (59.6% of 57)  - Etanercept: 127 males (74.7% of 170) - CZP 200 mg Q2W: 113 males (68.5% of 165) - CZP 400 mg Q2W: 107 males (64.1% of 167)  So the percentages of female participants can be calculated as:  - Placebo: 23 females (40.4% of 57) - Etanercept: 43 females (25.3% of 170)  - CZP 200 mg Q2W: 52 females (31.5% of 165) - CZP 400 mg Q2W: 60 females (35.9% of 167) |
| Mean PASI score at baseline | Certolizumab pegol 200 mg: 21.4 (8.8); Certolizumab pegol 400 mg: 20.8 (7.7); Etanercept: 21.0 (8.2); Placebo: 19.1 (17.1) | Table I of the manuscript provides the mean PASI score at baseline for each treatment group:  - Placebo: 19.1  - Etanercept: 21.0 - CZP 200 mg Q2W: 21.4 - CZP 400 mg Q2W: 20.8 |
| Mean duration of disease | Certolizumab pegol 200 mg: 19.5 (13.2); Certolizumab pegol 400 mg: 17.8 (11.5); Etanercept: 17.4 (12.0); Placebo: 18.9 (12.9) | Table I provides the mean duration of psoriasis at screening for each treatment group:  - Placebo: 18.9 years - Etanercept: 17.4 years - CZP 200 mg Q2W: 19.5 years  - CZP 400 mg Q2W: 17.8 years |
| Inclusion criteria | Adults with moderate-to-severe plaque psoriasis minimum 6 months’ duration, PASI ≥ 12, PGA ≥ 3 on a 5-point scale, and BSA ≥ 10%, candidates for systemic psoriasis therapy, phototherapy, or photochemotherapy. | The manuscript states the study inclusion criteria were:  - Adults with moderate-to-severe plaque psoriasis for ≥6 months  - Baseline PASI score ≥12  - Body surface area affected ≥10% - Baseline Physician's Global Assessment (PGA) score ≥3 on a 5-point scale - Candidates for systemic psoriasis therapy, phototherapy, or photochemotherapy |
| Exclusion criteria | Nonplaque psoriasis, history of recurrent infections or high risk for infection, malignancy, congestive heart failure, history of prior treatment with certolizumab pegol or etanercept, or failure of more than 2 biologic agents | The manuscript states the study exclusion criteria were:  - History of treatment with CZP, etanercept, or >2 biologic agents - History of primary failure to any biologic (no response within first 12 weeks) or secondary failure to >1 biologic (initial response then loss of response after week 12)  - Erythrodermic, guttate, or generalized pustular psoriasis - History of chronic or recurrent infections, including active or latent tuberculosis or high risk for infection - History of lymphoproliferative disorder, including lymphoma - History of malignancy or central nervous system demyelinating disease  - Congestive heart failure - Breastfeeding, pregnant, planning to become pregnant, or partner planning to become pregnant during study or within 3-5 months after last dose of study drug |
| N_randomized | 559 | According to the manuscript, a total of 559 patients were randomized in the study. |
| N_randomized_per_group | Certolizumab pegol 200 mg: 165; Certolizumab pegol 400 mg: 167; Etanercept: 170; Placebo: 57 | The manuscript states the randomization ratio was 3:3:1:3 for the treatment groups. Table I provides the number of participants randomized to each group:  - Placebo: 57 - Etanercept: 170  - CZP 200 mg Q2W: 165 - CZP 400 mg Q2W: 167 |
| N_analyzed | Certolizumab pegol 200 mg: 165; Certolizumab pegol 400 mg: 167; Etanercept: 170; Placebo: 57 | The manuscript does not explicitly state the number of participants analyzed in each treatment group. However, it provides some relevant information:  - Table II shows the number of participants in each group for the efficacy analyses (Placebo: 57, Etanercept: 170, CZP 200 mg Q2W: 165, CZP 400 mg Q2W: 167).   - Figure 2 shows the number of participants who completed week 16 in each initial treatment group (Placebo: 55, Etanercept: 161, CZP 200 mg Q2W: 156, CZP 400 mg Q2W: 161).  - Figure 2 also shows the number of PASI 75 responders at week 16 who entered the maintenance phase (Placebo: 2, Etanercept: 74, CZP 200 mg Q2W: 97, CZP 400 mg Q2W: 125).  So it seems the participant numbers analyzed for efficacy were similar to those originally randomized. |
| Dose_route_frequency | Certolizumab pegol 200 mg: 400 mg loading dose, subcutaneous, weeks 0, 2, and 4 then 200 mg every 2 weeks for 16 weeks; Certolizumab pegol 400 mg: 400 mg, subcutaneous, every 2 weeks for 16 weeks; Etanercept: 50 mg, subcutaneous, twice weekly for 12 weeks | According to the manuscript, the treatment interventions were:  - Placebo: administered subcutaneously every 2 weeks - Etanercept: 50 mg administered subcutaneously twice weekly  - CZP 200 mg: 200 mg subcutaneously every 2 weeks, with loading doses of 400 mg at weeks 0, 2, and 4 - CZP 400 mg: 400 mg subcutaneously every 2 weeks |
| Primary outcome_name | PASI 75 | The primary efficacy endpoint was PASI 75 ($75% reduction in Psoriasis Area and Severity Index from baseline PASI) responder rate for both CZP doses versus placebo at week 12. |
| Primary outcome_estimate | Certolizumab pegol 200 mg: 101 (61.3);  Certolizumab pegol 400 mg: 111 (66.7);  Etanercept: 91 (53.3);  Certolizumab pegol 200 mg vs. etanercept: ARD=8.0 (95% CI, –2.9 to 18.9); OR=1.4 (95% CI, 0.9 to 2.2), p=0.15; RR=1.1 (95% CI, 0.95 to 1.4), p=0.16;   Certolizumab pegol 400 mg vs. etanercept: ARD=13.4 (95% CI, 2.7 to 24.1); OR=1.8 (95% CI, 1.1 to 2.8), p=0.02; RR=1.2 (95% CI, 1.04 to 1.5), p=0.02 | Here are the effect estimates for the primary outcome of PASI 75 response at week 12:  - Placebo group: PASI 75 response in 5.0% of patients  - CZP 200 mg Q2W group: PASI 75 response in 61.3% of patients. Odds ratio vs placebo 30.0 (95% CI 9.0 to 100.5), p<0.0001  - CZP 400 mg Q2W group: PASI 75 response in 66.7% of patients. Odds ratio vs placebo 38.0 (95% CI 11.3 to 127.6), p<0.0001  - Etanercept group: PASI 75 response in 53.3% of patients  - CZP 400 mg vs Etanercept:   - Difference in PASI 75 response: 13.4% (95% CI 2.7% to 24.1%)  - Odds ratio: 1.8 (95% CI 1.1 to 2.8), p=0.0152  - CZP 200 mg vs Etanercept:  - Difference in PASI 75 response: 8.0% (95% CI -2.9% to 18.9%)  - Odds ratio: 1.4 (95% CI 0.9 to 2.2)   - Met criteria for non-inferiority  So in summary, both CZP doses showed significantly higher PASI 75 response versus placebo, CZP 400 mg was superior to etanercept, and CZP 200 mg was non-inferior to etanercept. |

**Papp et al. 2018^10^**

| **Variable** | **Reference standard** | **Claude 2^9^** |
| --- | --- | --- |
| First author's last name | Papp | The last name of the first author is Papp. |
| Trial registry number | NCT02905006 | The registry number of the study is NCT02905006, which is stated in the Methods section:  "This randomized, double-blinded, placebo-controlled, parallel group, dose-ranging study (NCT02905006) recruited patients during August 25, 2016-March 1, 2017, in 6 countries (Canada, Czech Republic, Hungary, Japan, Poland, and United States)." |
| Study name and acronym | BE ABLE 1 | The trial name is not explicitly stated in the paper. However, based on the title "Dual neutralization of both interleukin 17A and interleukin 17F with bimekizumab in patients with psoriasis: Results from BE ABLE 1, a 12-week randomized, double-blinded, placebo-controlled phase 2b trial", it appears the trial name is "BE ABLE 1". |
| Study funder | UCB Pharma | UCB Pharma |
| Mean age | 44.3 (13.7) | Based on Table 1, the mean age in years with standard deviation reported to one decimal place for each treatment group and overall was:  - Placebo: 46.7 (12.3) - BKZ 64 mg: 44.2 (13.8)  - BKZ 160 mg: 43.4 (12.4) - BKZ 160 mg (320 mg LD): 46.5 (15.2) - BKZ 320 mg: 42.6 (13.6) - BKZ 480 mg: 42.9 (15.2) - All patients: 44.3 (13.7) |
| Female participants | 87 (34.8) | According to Table 1, the total count and corresponding percentage of female participants rounded to one decimal place in each treatment group and overall were:  - Placebo: 17, 40.5% - BKZ 64 mg: 19, 48.7%  - BKZ 160 mg: 11, 25.6% - BKZ 160 mg (320 mg LD): 11, 27.5% - BKZ 320 mg: 15, 34.9% - BKZ 480 mg: 14, 32.6% - Total: 87, 34.8% |
| Mean PASI score at baseline | 19.1 (6.5) | Based on Table 1, the mean PASI score at baseline with standard deviation for each treatment group and total was:  - Placebo: 18.9 (5.8) - BKZ 64 mg: 18.6 (5.9) - BKZ 160 mg: 20.6 (6.8)  - BKZ 160 mg (320 mg LD): 18.9 (8.4) - BKZ 320 mg: 19.4 (6.4) - BKZ 480 mg: 18.4 (5.1) - All patients: 19.1 (6.5) |
| Mean duration of disease | 15.0 (0-58.7) | The median number of years that participants had plaque psoriasis for each treatment group, reported to one decimal place, was:  - Placebo: 15.0 - BKZ 64 mg: 15.0 - BKZ 160 mg: 15.9 - BKZ 160 mg (320 mg LD): 14.5  - BKZ 320 mg: 15.9 - BKZ 480 mg: 13.9  This information is provided in Table 1 under the "Disease duration, years, median (range)" row. |
| Inclusion criteria | Adults with moderate-to-severe plaque psoriasis minimum 6 months’ duration, ≥ 10% BSA, IGA score of ≥ 3 on a 5-point scale, and candidate for systemic psoriasis therapy or phototherapy. | The study inclusion criteria were:  - Patients were at least 18 years of age. - They had chronic plaque psoriasis for at least 6 months before screening.  - They had a Psoriasis Area and Severity Index (PASI) ≥12 and body surface area (BSA) ≥10%, and Investigator’s Global Assessment (IGA) score on a 5-point scale of ≥3. - They were candidates for systemic psoriasis therapy, phototherapy, or chemophototherapy.  - Female patients were postmenopausal, permanently sterilized, or (if of childbearing potential) were willing to use a highly effective method of contraception until 20 weeks after last administration of study medication and had a negative pregnancy test at visit 1 (screening) and immediately before first dose.  - Male patients with a partner of childbearing potential used a condom when sexually active, until 20 weeks after the last administration of study medication. - Patients agreed to not increase their usual sun exposure during the study and to use sunscreens that protected against ultraviolet A (UVA) and ultraviolet B (UVB).  These are listed in the Inclusion Criteria section of the Supplementary Appendix. |
| Exclusion criteria | Prior treatment with an anti-IL-17 therapy or prior exposure to > 1 other biologic therapy for psoriasis or psoriatic arthritis, a significant uncontrolled neuropsychiatric disorder, history of a suicide attempt, or suicide ideation within 6 months. | The study exclusion criteria were:  - Patients were female patients who were breastfeeding, pregnant, or planned to become pregnant during the study or within 20 weeks after last dose of study drug. Male patients were planning to impregnate their partners during the study or within 20 weeks after the last dose. - They previously participated in a bimekizumab clinical trial or another study of a medication or medical device under investigation within the past 3 months or at least 5 half-lives, whichever was greater, or were currently participating in another study of a medication or medical device under investigation.  - They had a known hypersensitivity to any excipients of bimekizumab. - They had an erythrodermic, guttate, pustular form of psoriasis or drug-induced psoriasis. - They had a history of chronic or recurrent infections, or a serious or life-threatening infection within the 6 months before the baseline visit. - And several other criteria listed in the Exclusion Criteria section of the Supplementary Appendix. |
| N_randomized | 250 | The total number of participants who were randomly assigned to any treatment group in the study was 250. This is stated in the Results section:  "In this study, 355 patients were screened, with 250 randomized to treatment (Supplemental Figs 1 and 2; available at http://www.jaad.org)." |
| N_randomized_per_group | Placebo: 42; Bimekizumab 64 mg: 39; Bimekizumab 160 mg: 43; Bimekizumab 160 mg (320 mg at baseline): 40; Bimekizumab 320 mg: 43; Bimekizumab 480 mg: 43 | The number of participants randomly assigned to each group was:  - Placebo: 42 - BKZ 64 mg: 39  - BKZ 160 mg: 43 - BKZ 160 mg (320 mg LD): 40 - BKZ 320 mg: 43 - BKZ 480 mg: 43  This information is provided in the CONSORT flow diagram in Supplemental Figure 2. |
| N_analyzed | Bimekizumab 64 mg: 39; Bimekizumab 160 mg: 43; Bimekizumab 160 mg (320 mg at baseline): 40; Bimekizumab 320 mg: 43; Bimekizumab 480 mg: 43 | The number of participants analyzed in each group (the full analysis set) was:  - Placebo: 42  - BKZ 64 mg: 39 - BKZ 160 mg: 43 - BKZ 160 mg (320 mg LD): 40 - BKZ 320 mg: 43 - BKZ 480 mg: 43  This is stated in the Results section:  "All randomized patients were included in the full analysis set."  And also shown in the CONSORT flow diagram in Supplemental Figure 2. |
| Dose_route_frequency | Bimekizumab: 64 mg, 160 mg, 160 mg (with 320 mg loading dose at baseline), 320 mg, 480 mg, subcutaneous, week 0, week 4, week 8 | The dose, route of administration, and frequency for each treatment were:  - Placebo: 0 mg subcutaneous injection every 4 weeks - BKZ 64 mg: 64 mg subcutaneous injection every 4 weeks - BKZ 160 mg: 160 mg subcutaneous injection every 4 weeks  - BKZ 160 mg (320 mg LD): 160 mg subcutaneous injection every 4 weeks (with 320 mg loading dose at baseline) - BKZ 320 mg: 320 mg subcutaneous injection every 4 weeks - BKZ 480 mg: 480 mg subcutaneous injection every 4 weeks  This information is provided in the Methods section:  "Patients were randomized (1:1:1:1:1:1) to receive bimekizumab every 4 weeks at doses of 64 mg, 160 mg, 160 mg (with 320 mg loading dose at baseline), 320 mg, 480 mg, or placebo."  And:  "Bimekizumab was provided in single-use vials containing 160 mg/mL. Due to differences in presentation and to ensure study blinding, bimekizumab and placebo injections were prepared and administered at the investigational sites by unblinded, dedicated study personnel." |
| Primary outcome_name | PASI 90 | The primary outcome designated by the study authors was PASI90 response at week 12.   This is stated in the Methods section:  "The primary efficacy endpoint was PASI90 at week 12."  Where PASI90 is defined as "$90% reduction in Psoriasis Area and Severity Index (PASI) from baseline readings" |
| Primary outcome_estimate | Bimekizumab 64 mg: 46.2; Bimekizumab 160 mg: 67.4; Bimekizumab 160 mg (320 mg at baseline): 75.0; Bimekizumab 320 mg: 79.1; Bimekizumab 480 mg: 72.1; Placebo: 0%; p< 0.001, all comparisons | The primary outcome (PASI90 response at week 12) was a dichotomous outcome reported as counts and proportions. The results for each treatment group were:  - Placebo: 0/42 (0%) - BKZ 64 mg: 18/39 (46.2%), p<.0001 vs placebo - BKZ 160 mg: 27/43 (62.8%), p<.0001 vs placebo  - BKZ 160 mg (320 mg LD): 30/40 (75.0%), p<.0001 vs placebo - BKZ 320 mg: 34/43 (79.1%), p<.0001 vs placebo - BKZ 480 mg: 29/43 (67.4%), p<.0001 vs placebo  This is presented in Fig. 3 and stated in the Results section:  "PASI90 response at week 12 was achieved by significantly more patients in all bimekizumab-treatment groups compared with the placebo group (46.2%-79.1% vs 0%; P < .0001, all comparisons)" |

**Reich et al. 2017 (LIBERATE)^5^**

| **Variable** | **Reference standard** | **Claude 2^9^** |
| --- | --- | --- |
| First author's last name | Reich | The last name of the first author is Reich. |
| Trial registry number | NCT01690299 | The clinicaltrials.gov identifier for this study is NCT01690299. This is stated in the Methods section of the paper:  "LIBERATE was a global, phase IIIb, multi-centre, randomized, double-blind, placebo-controlled study (NCT01690299)." |
| Study name and acronym | LIBERATE | The trial name is "Evaluation in a Placebo-Controlled Study of Oral Apremilast and Etanercept in Plaque Psoriasis".   The acronym for the trial name is LIBERATE. This is mentioned in the first paragraph of the Introduction section:  "Here, we report the findings from a phase IIIb study (Evaluation in a Placebo-Controlled Study of Oral Apremilast and Etanercept in Plaque Psoriasis [LIBERATE]) that evaluated apremilast vs. placebo at Week 16 in biologic-naive patients with moderate-to-severe plaque psoriasis." |
| Study funder | Celgene | The study was sponsored by Celgene Corporation. This is stated in the Funding sources section:  "This study was sponsored by Celgene Corporation." |
| Mean age | Placebo: 43.4 (14.9); Apremilast: 46.0 (13.6); Etanercept: 47.0 (14.1) | According to Table 1, the mean age and standard deviation for each treatment group was:   Placebo: 43.4 years (SD 14.9 years)  Apremilast: 46.0 years (SD 13.6 years)  Etanercept: 47.0 years (SD 14.1 years)  Reporting the mean and standard deviation to one decimal place:   Placebo: 43.4 years (SD 14.9 years)  Apremilast: 46.0 years (SD 13.6 years)  Etanercept: 47.0 years (SD 14.1 years) |
| Female participants | Placebo: 25 (29.8); Apermilast: 34 (41.0); Etanercept: 34 (41.0) | The total counts and percentages of female participants in each treatment group were:  - Placebo: 25 out of 84 (29.8%)  - Apremilast: 34 out of 83 (41.0%) - Etanercept: 34 out of 83 (41.0%)  These counts can be derived from Table 1, which reports the number and percentage of male participants in each group. By subtracting the number of males from the total number of participants in each group, the number of females can be determined. |
| Mean PASI score at baseline | Placebo: 19.4 (6.8) Apremilast: 19.3 (7.0) Etanercept: 20.3 (7.9) | According to Table 1 in the paper, the mean PASI score and standard deviation at baseline for each treatment group was:  - Placebo: 19.4 (SD 6.8) - Apremilast: 19.3 (SD 7.0)  - Etanercept: 20.3 (SD 7.9)  To report this to one decimal place:  - Placebo: Mean PASI 19.4 (SD 6.8) - Apremilast: Mean PASI 19.3 (SD 7.0) - Etanercept: Mean PASI 20.3 (SD 7.9) |
| Mean duration of disease | Placebo: 16.6 (12.1); Apremilast: 19.7 (12.7); Etanercept: 18.1 (11.7) | According to Table 1 in the paper, the mean duration of psoriasis in years and standard deviation for each treatment group was:  - Placebo: 16.6 years (SD 12.1 years) - Apremilast: 19.7 years (SD 12.7 years) - Etanercept: 18.1 years (SD 11.7 years)  Reporting this to one decimal place:  - Placebo: 16.6 years (SD 12.1 years) - Apremilast: 19.7 years (SD 12.7 years)  - Etanercept: 18.1 years (SD 11.7 years) |
| Inclusion criteria | Adults 18 years and older with chronic plaque psoriasis for at least 12 months (PASI score ≥12, BSA ≥10%, sPGA score ≥3 ), inadequate response, intolerance or contraindication to at least 1 conventional systemic agent for treatment of psoriasis; candidate for phototherapy or systemic therapy | "The key inclusion criteria were:  - Adults aged ≥18 years  - Chronic plaque psoriasis for ≥12 months  - PASI score ≥12, affected body surface area ≥10%, static Physician Global Assessment score ≥3 (moderate to severe)  - Inadequate response, intolerance or contraindication to ≥1 conventional systemic agent for psoriasis  - Candidates for phototherapy or systemic (including etanercept) therapy  - No prior exposure to a biologic therapy for psoriasis or psoriatic arthritis  These criteria are listed in the Study population section of the Methods." |
| Exclusion criteria | * Prior exposure to a biologic therapy,  * prior failure of more than 3 systemic agents for treatment of psoriasis,  * history of known demyelinating diseases such as multiple sclerosis or optic neuritis,  * history of or concurrent congestive heart failure, including medically controlled, asymptomatic congestive heart failure,  * other clinically significant or major uncontrolled disease, serious infection, latent, active or history of incompletely treated tuberculosis | Here is the complete list of exclusion criteria from the study as a bulleted list:  - Prior failure of >3 systemic agents for treatment of psoriasis - History of known demyelinating diseases such as multiple sclerosis or optic neuritis  - History of or concurrent congestive heart failure, including medically controlled, asymptomatic congestive heart failure - Other clinically significant or major uncontrolled disease - Serious infection  - Latent, active or history of incompletely treated tuberculosis - Malignancy or history of malignancy, except successfully treated basal cell carcinoma or squamous cell carcinoma of the skin or carcinoma in situ of the cervix - Laboratory abnormalities including:  - Positive urine drug screen  - Positive hepatitis B surface antigen or hepatitis C antibody  - Abnormal liver function tests (ALT, AST >1.5x ULN)  - Abnormal renal function (creatinine >1.7 mg/dL) - Live vaccination within 4 weeks prior to baseline - Requirement for >2 topical corticosteroids for scalp psoriasis - Previous use of biologic therapy for psoriasis or psoriatic arthritis - Prior use of apremilast - Pregnant or breastfeeding women |
| N_randomized | 250 | A total of 250 patients were randomized in the study. This is stated in the Results section:  "A total of 250 patients were randomized and included in the full analysis set (placebo, n = 84; apremilast, n = 83; etanercept, n = 83)." |
| N_randomized_per_group | Apremilast: 83; Etanercept: 83; Placebo: 84 | The number of participants randomly assigned to each treatment group was:  - Placebo: 84 - Apremilast: 83  - Etanercept: 83  This is reported in the Results section:  "A total of 250 patients were randomized and included in the full analysis set (placebo, n = 84; apremilast, n = 83; etanercept, n = 83)." |
| N_analyzed | Apremilast: 83; Etanercept: 83; Placebo: 84  (used a mITT and therefore, all randomized were included in the analysis; however, the number of individuals who completed the trial: Placebo: 75; Apremilast: 77; Etanercept: 81) | The number of participants analyzed in each treatment group (the modified intent-to-treat population) was:  - Placebo: 84  - Apremilast: 83 - Etanercept: 83  This is stated in the Statistical analysis section of Methods:  "Efficacy assessments were conducted for the modified intent-to-treat (mITT) population (all randomized patients who received ≥1 dose of study medication and had both baseline PASI and ≥1 post-treatment PASI evaluations)."  And confirmed by the Results section stating 250 patients were randomized and included in the full analysis set, with n=84, 83, and 83 for each group. |
| Dose_route_frequency | Apremilast: 30 mg, oral, twice per day (BID); Etanercept: 50 mg, subcutaneous injection, once weekly | The interventions and dosing were:  - Placebo: placebo tablets orally twice daily + placebo subcutaneous injections once weekly - Apremilast: 30 mg tablets orally twice daily + placebo subcutaneous injections once weekly  - Etanercept: placebo tablets orally twice daily + 50 mg subcutaneous injections once weekly  This information is provided in the Study design section of Methods:  "Eligible patients were randomized (1:1:1) via an interactive voice response system to placebo; apremilast oral tablet, 30 mg BID; or etanercept subcutaneous injection, 50 mg QW (placebo-controlled phase; Fig. 1)." |
| Primary outcome_name | PASI 75 | The primary efficacy endpoint was the proportion of patients who achieved a ≥75% reduction from baseline in Psoriasis Area and Severity Index (PASI-75) score at Week 16 with apremilast compared to placebo.  This is stated in the Assessments section of Methods:  "The primary efficacy endpoint was the proportion of patients who achieved a ≥75% reduction from baseline in Psoriasis Area and Severity Index score (PASI-75) at Week 16 with apremilast or placebo." |
| Primary outcome_estimate | Placebo: 11.9%; Apremilast: 39.8%; Etanercept: 48.2%; Apremilast vs. Etanercept: p=0.26  (The comparison of PASI 75 for apremilast vs. etanercept is a post-hoc analysis; these data are based on LOCF; authors also report primary endpoint using non-responder imputation) | Here are the effect estimates for the primary outcome PASI-75 response at Week 16 for each treatment group and treatment comparisons:  - Apremilast:   - 39.8% (33/83 patients) achieved PASI-75  - Placebo:  - 11.9% (10/84 patients) achieved PASI-75   - Etanercept:  - 48.2% (40/83 patients) achieved PASI-75  Treatment comparisons:  - Apremilast vs Placebo  - Absolute risk reduction: 39.8% - 11.9% = 27.9%   - Relative risk: 39.8% / 11.9% = 3.34  - Odds ratio: not reported  - P<0.0001  - Etanercept vs Placebo  - Absolute risk reduction: 48.2% - 11.9% = 36.3%  - Relative risk: 48.2% / 11.9% = 4.05  - Odds ratio: not reported  - P<0.0001  - Apremilast vs Etanercept  - Absolute risk reduction: not reported  - Relative risk: not reported  - Odds ratio: not reported   - P-value: not reported (study was not powered for this comparison)  In summary, the study reported proportions and p-values, but did not provide relative risks, odds ratios or other effect measures for the treatment comparisons. I calculated the absolute risk reductions and relative risks based on the proportions provided. |

**Reich et al. 2017 (VOYAGE)^6^**

| **Variable** | **Reference standard** | **Claude 2^9^** |
| --- | --- | --- |
| First author's last name | Reich | The first author's last name is Reich. |
| Trial registry number | NCT02207244 | The registry numbers for this study are not listed in the paper. The paper mentions that it is part of the VOYAGE clinical trial program, but does not provide specific registry numbers. |
| Study name and acronym | VOYAGE-2 | The trial name is "VOYAGE 2". It does not appear to have an acronym specified in the paper. |
| Study funder | Jannsen Research & Development | The paper states that the study was supported by Janssen Research & Development, LLC. |
|  | Adalimumab: 43.2 (11.9); Guselkumab: 43.7 (12.2) | According to Table II in the paper, the average age and standard deviation for each treatment group was:  Placebo: 43.3 ± 12.4 years Guselkumab: 43.7 ± 12.2 years  Adalimumab: 43.2 ± 11.9 years  Reporting the mean and standard deviation to one decimal place:  Placebo: 43.3 ± 12.4 years Guselkumab: 43.7 ± 12.2 years Adalimumab: 43.2 ± 11.9 years |
| Female participants | Adalimumab: 78 (31.5); Guselkinumab: 147 (29.6) | According to Table II of the study, the number and percentage of female participants in each treatment group was:  Placebo:  - Total patients: 248 - Female patients: 75  - Percentage female: 30.2%  Guselkumab: - Total patients: 496  - Female patients: 147 - Percentage female: 29.6%  Adalimumab: - Total patients: 248 - Female patients: 78 - Percentage female: 31.5%  Rounding the percentages to one decimal place:  Placebo: 30.2% female  Guselkumab: 29.6% female Adalimumab: 31.5% female |
| Mean PASI score at baseline | Adalimumab: 21.7 (9.0); Guselkumab: 21.9 (8.8) | According to Table II of the study, the mean PASI scores at baseline for each treatment group were:  Placebo: 21.5 Guselkumab: 21.9  Adalimumab: 21.7  Therefore, the mean PASI scores at baseline were:  Placebo: 21.5 Guselkumab: 21.9  Adalimumab: 21.7 |
| Mean duration of disease | Adalimumab: 17.6 (11.7); Guselkumab: 17.9 (12.0) | According to Table II of the study, the average number of years patients had plaque psoriasis in each treatment group was:  Placebo: 17.9 years Guselkumab: 17.9 years Adalimumab: 17.6 years  Reporting the average duration of psoriasis to one decimal place:  Placebo: 17.9 years Guselkumab: 17.9 years  Adalimumab: 17.6 years |
| Inclusion criteria | Adults aged 18 and older with moderate-to-severe plaque psoriasis, IGA score ≥ 3, PASI score ≥ 12, BSA ≥ 10% for at least 6 months and candidates for systemic therapy or phototherapy. | The main inclusion criteria listed for participants in this study were:  - Adults aged ≥18 years  - Diagnosis of moderate to severe plaque psoriasis  No other specific inclusion criteria were mentioned in the provided text. |
| Exclusion criteria | History or current signs of a severe, progressive, or uncontrolled medical condition or current or history of malignancy, except nonmelanoma skin cancer, within 5 years, previously received guselkumab or adalimumab, other anti-TNF-α therapy within 3 months, other treatment targeting IL-12/23, IL-17, or IL-23 in prior 6 months, any systemic immunosuppressants (e.g., methotrexate) or phototherapy in prior 4 weeks. | The study exclusion criteria are not explicitly stated in the provided text. The text mentions "Major inclusion/exclusion criteria are summarized in VOYAGE 1", but does not provide the specific exclusion criteria for this study. |
| N_randomized | 922 | The total number of participants randomized to any treatment group in the study was 992. This included:  - Placebo: 248 - Guselkumab: 496 - Adalimumab: 248 |
| N_randomized_per_group | Placebo: 248; Adalimumab: 248; Guselkumab: 496 | The number of participants randomly assigned to each group was:  - Placebo: 248  - Guselkumab: 496 - Adalimumab: 248 |
| N_analyzed | Placebo: 248; Adalimumab: 248; Guselkumab: 496 | The number of participants analyzed in each group is not explicitly stated in the provided text. The results sections mention analysis of the randomized patients, but do not provide counts of how many were analyzed in each group. |
| Dose_route_frequency | Adalimumab: 80 mg, subcutaneous, week 0, 40 mg, subcutaneous, week 1 and every 2 weeks; Guselkumab: 100 mg, subcutaneous, weeks 0, 4, 12 | The dose, route of administration, and frequency for each treatment were:  - Placebo: Not applicable  - Guselkumab: 100 mg subcutaneous injection at weeks 0, 4, then every 8 weeks - Adalimumab: 80 mg subcutaneous injection at week 0, then 40 mg subcutaneous injection at week 1 and every 2 weeks through week 23 |
| Primary outcome_name | IGA 0/1 at week 16; PASI 90 at week 16 | The coprimary endpoints of the study were:  - Proportion of patients achieving an Investigator Global Assessment (IGA) score of cleared/minimal (0/1) at week 16 - Proportion of patients achieving a 90% or greater improvement in Psoriasis Area and Severity Index (PASI 90) response at week 16 |
| Primary outcome_estimate | IGA 0 or 1: Guselkumab: 417 (84.1); Adalimumab: 168 (67.7); ARD=16.3% (95% CI, 9.7% to 23.0%); RR=1.2 (95% CI, 1.1 to 1.4); PASI 90: Guselkumab: 347 (70.0); Adalimumab: 116 (46.8); ARD=23.2% (95% CI, 15.8% to 30.6%); RR=1.5 (95% CI, 1.3 to 1.7)  (for guselkumab vs placebo. See table 1 of 36519. no primary outcomes for guselkumab vs adalimumab.   Risk ratio and risk difference calculated) | The primary outcome results for each treatment group were:  - Placebo:   - IGA 0/1 at week 16: 21/248 (8.5%)  - PASI 90 at week 16: 6/248 (2.4%)  - Guselkumab:  - IGA 0/1 at week 16: 417/496 (84.1%)   - PASI 90 at week 16: 347/496 (70.0%)  The comparisons between guselkumab and placebo for the coprimary endpoints were statistically significant (p < 0.001). |

**Reich et al. 2017 (IXORA-S)^7^**

| **Variable** | **Reference standard** | **Claude 2^9^** |
| --- | --- | --- |
| First author's last name | Reich | The last name of the first author is Reich. |
| Trial registry number | NCT02561806 | The clinical trial registration number for this study is NCT02561806, as stated in the Methods section:  "This 52-week, phase IIIb, multicentre, controlled, double-blind, parallel-group trial (IXORA-S, NCT02561806) was conducted at 51 sites across 13 countries." |
| Study name and acronym | IXORA-S | The trial name is "Comparison of ixekizumab with ustekinumab in moderate-to-severe psoriasis: 24-week results from IXORA-S, a phase III study".   The acronym for the trial name is IXORA-S. |
| Study funder | Eli Lilly and Company | The study was funded in full by Eli Lilly and Company, as stated in the Funding Sources section:  "This study was funded in full by Eli Lilly and Company, Indianapolis, IN, U.S.A." |
| Mean age | Ixekizumab: 42.7 (12.7); Ustekinumab: 44.0 (13.3) | According to Table 1 in the paper, the average age and standard deviation of study participants in each treatment group was:  - Ustekinumab group: 44.0 ± 13.3 years - Ixekizumab group: 42.7 ± 12.7 years  Reporting the averages to one decimal place:  - Ustekinumab group: 44.0 ± 13.3 years - Ixekizumab group: 42.7 ± 12.7 years |
| Female participants | Ixekizumab: 46 (33.8); Ustekinumab: 54 (32.5) | The number and percentage of female participants in each treatment group is:  - Ustekinumab group: 54 (32.5%)  - Ixekizumab group: 46 (33.8%)  As stated in the Baseline demographics and clinical characteristics table:  "Sex (male), n (%) Ustekinumab: 112 (67.5)  Ixekizumab: 90 (66.2)"  So the number of females can be calculated as:  - Ustekinumab group: 166 total - 112 males = 54 females (32.5%) - Ixekizumab group: 136 total - 90 males = 46 females (33.8%) |
| Mean PASI score at baseline | Ixekizumab: 19.9 (8.2); Ustekinumab: 19.8 (9.0) | According to Table 1, the mean PASI score with standard deviation at baseline for each treatment group was:  - Ustekinumab group: 19.8 ± 9.0  - Ixekizumab group: 19.9 ± 8.2 |
| Mean duration of disease | Ixekizumab: 18.0 (1.1); Ustekinumab: 18.2 (12.0) | According to Table 1, the average number of years with standard deviation that participants had plaque psoriasis for each treatment group was:  - Ustekinumab group: 18.2 ± 12.0 years - Ixekizumab group: 18.0 ± 11.1 years   Reporting the averages to one decimal place:  - Ustekinumab group: 18.2 ± 12.0 years - Ixekizumab group: 18.0 ± 11.1 years |
| Inclusion criteria | Adults aged 18 and older with moderate-to-severe plaque psoriasis minimum 6 months’ duration and PASI ≥ 10, previously failed or had a contraindication or intolerability to at least 1 systemic therapy (including ciclosporin, methotrexate and phototherapy). | The key inclusion criteria for the study are stated in the Study population section:  "Eligible study participants were aged ≥ 18 years, had a diagnosis of chronic plaque psoriasis for ≥ 6 months, had a PASI score ≥ 10 and had previously failed or had a contraindication or intolerability to at least one systemic therapy (including ciclosporin, methotrexate and phototherapy)." |
| Exclusion criteria | Predominant presence of nonplaque psoriasis, a contraindication for ustekinumab, or prior treatment with ustekinumab, ixekizumab or any other IL-17 or IL-12/23 | The key exclusion criteria are stated in the Study population section:  "Key exclusion criteria were a predominant presence of nonplaque psoriasis, a contraindication for ustekinumab, or prior treatment with ustekinumab, ixekizumab or any other IL-17 or IL-12/23 antagonists." |
| N_randomized | 302 | The total number of participants randomly assigned to a treatment group is 302, as stated in the Study population section:  "In total, 355 patients were screened (Fig. 2), of whom 302 were randomized to receive ustekinumab (n = 166) or ixekizumab (n = 136)." |
| N_randomized_per_group | Ixekizumab: 136; Ustekinumab: 166 | The number of participants randomly assigned to each treatment group is:  - Ustekinumab group: 166  - Ixekizumab group: 136  As stated in the Study population section:  "In total, 355 patients were screened (Fig. 2), of whom 302 were randomized to receive ustekinumab (n = 166) or ixekizumab (n = 136)." |
| N_analyzed | Ixekizumab: 136; Ustekinumab: 166 | The number of participants analyzed in each treatment group is:  - Ustekinumab group: 166 - Ixekizumab group: 136  As stated in the Statistical analyses section:  "Patients were analysed according to the treatment they were assigned at randomization (intention-to-treat population)."  And the participant counts match those originally randomized as stated in the Study population section. |
| Dose_route_frequency | Ixekizumab: 80 mg, subcutaneous, every 2 weeks through week 12, then every 4 weeks; Ustekinumab: 45 or 90 mg (if patient weight more than 100 kg), subcutaneous, weeks 0, 4, and 16 | The dose, route of administration, and frequency for each treatment is:  Ustekinumab: - Dose: 45 mg if ≤100 kg, 90 mg if >100 kg  - Route: Subcutaneous injection - Frequency: Weeks 0, 4, 16, 28, 40  Ixekizumab: - Dose: 160 mg at week 0, then 80 mg  - Route: Subcutaneous injection  - Frequency: Every 2 weeks through week 12, then every 4 weeks  As stated in the Study design section:  "Patients randomized to ixekizumab received two subcutaneous (SC) injections of ixekizumab 80 mg (160 mg total) at week 0, followed by one SC injection of ixekizumab 80 mg every 2 weeks through week 12, and 80 mg every 4 weeks thereafter."  "Patients randomized to ustekinumab were dosed at weeks 0, 4, 16, 28 and 40, in accordance with the label, with patients weighing ≤ 100 kg receiving 45 mg SC injections and patients weighing > 100 kg receiving 90 mg SC injections." |
| Primary outcome_name | PASI 90 | The primary outcome of the study was the proportion of patients achieving ≥ 90% improvement in Psoriasis Area and Severity Index (PASI 90) at week 12.  As stated in the Study objectives section:  "The primary objective of IXORA-S was to demonstrate firstly that ixekizumab is noninferior to ustekinumab (inferiority margin: −12.6%) and secondly that ixekizumab is superior to ustekinumab, as measured by the proportion of patients achieving a PASI 90 response at week 12." |
| Primary outcome_estimate | Ixekizumab: 72.8%; Ustekinumab: 42.2%, ARD=32.1%, (97.5% CI, 19.8 to 44.5%), p<0.001 | For the primary outcome of PASI 90 response at week 12:  - Ixekizumab group: 99/136 patients (72.8%) - Ustekinumab group: 70/166 patients (42.2%)   The response difference was 32.1% in favor of ixekizumab (97.5% CI 19.8–44.5%, p<0.001).  As stated in the Primary end point: week 12 section:  "At week 12, significantly more patients in the ixekizumab group (n = 99, 72.8%) than in the ustekinumab group (n = 70, 42.2%) achieved PASI 90 (response difference 32.1%, 97.5% confidence interval 19.8–44.5%, P < 0.001)." |

**Thaçi et al. 2015^8^**

| **Variable** | **Reference standard** | **Claude 2^9^** |
| --- | --- | --- |
| First author's last name | Thaci | Thaçi |
| Trial registry number | NCT02074982 | The clinical trials registry number for this study is NCT02074982. This is stated in the Methods section:  "This 52-week, randomized, double-blind, active comparator, parallel-group, superiority phase IIIb study was conducted in accordance with ethical principles of the Declaration of Helsinki at 134 sites worldwide. US sites maintained compliance with Health Insurance Portability and Accountability Act regulations. The study was initiated in February 2014 (first subject, first visit), and last subject, last visit for the week-16 primary efficacy analysis occurred in October 2014." |
| Study name and acronym | CLEAR | The trial name is "A Randomized Head-to-Head Study to Evaluate The Efficacy and Safety of Secukinumab Compared to Ustekinumab in Patients With Moderate to Severe Plaque-Type Psoriasis".  The acronym for the trial name is CLEAR, which stands for "CLear skin Efficacy Assessment using secukinuMab vs. ustekinumab". This is mentioned in the first paragraph of the introduction:  "CLEAR, the second head-to-head trial of secukinumab, directly compared the efficacy and safety of secukinumab with ustekinumab in subjects with moderate to severe plaque psoriasis." |
| Study funder | Novartis Pharma AG | The study was funded and sponsored by Novartis Pharma AG, which is stated in the disclosures section:  "Novartis Pharma AG (Basel, Switzerland) supported this study." |
| Mean age | Secukinumab: 45.2 (13.96); Ustekinumab: 44.6 (13.67) | According to Table I in the paper, the average age with standard deviation for each treatment group was:  - Secukinumab group: 45.16 ± 13.96 years - Ustekinumab group: 44.64 ± 13.67 years  The average ages are reported to two decimal places as requested. |
| Female participants | Secukinumab: 108 (32.0);  Ustekinumab: 87 (25.7) | The number and percentage of female participants in each treatment group is:  Secukinumab group:  - Total participants: 337 - Female participants: 108 (32.0%)  Ustekinumab group: - Total participants: 339  - Female participants: 87 (25.7%)  This is based on the data reported in Table 1:  "Male gender 229 (68.0) 252 (74.3)"  So the female percentages can be calculated as:  Secukinumab: 337 total - 229 males = 108 females → 108/337 = 32.0% female Ustekinumab: 339 total - 252 males = 87 females → 87/339 = 25.7% female |
| Mean PASI score at baseline | Secukinumab: 21.7 (8.50); Ustekinumab: 21.5 (8.07) | According to Table 1 of the study, the mean PASI score with standard deviation at baseline for each treatment group was:   Secukinumab 300 mg: 21.7 ± 8.5  Ustekinumab: 21.5 ± 8.1 |
| Mean duration of disease | Secukinumab: 19.6 (12.90); Ustekinumab: 16.1 (11.24) | According to Table 1 of the study, the average number of years with standard deviation that participants had plaque psoriasis for each treatment group was:  - Secukinumab 300 mg: 19.6 ± 12.9 years - Ustekinumab: 16.1 ± 11.2 years  Reporting the averages to one decimal place:  - Secukinumab 300 mg: 19.6 ± 12.9 years  - Ustekinumab: 16.1 ± 11.2 years |
| Inclusion criteria | Adults aged 18 and older with moderate-to-severe plaque psoriasis for a minimum of 6 months’ duration and ≥ 10% BSA, PASI ≥ 12, IGA 2011 modified version 3 (moderate) or 4 (severe) and inadequate response to topical treatment, and/or phototherapy, and/or previous systemic therapy (conventional or biologic). | - Adults aged ≥18 years - Diagnosis of moderate to severe plaque psoriasis at least 6 months prior to enrollment  - Inadequate disease control with topical treatments, phototherapy, and/or previous systemic therapies |
| Exclusion criteria | Prior exposure to any biologics directly targeting IL-17A/IL-17 receptor A or IL-12/IL-23. | The key exclusion criteria are not comprehensively listed in the article, but the Methods section states:  "Key exclusion criteria included previous exposure to any biologics directly targeting IL-17A/IL-17 receptor A or IL-12/IL-23."  So the key exclusion criterion was:  - Previous exposure to biologics that directly target IL-17A/IL-17 receptor A (e.g. secukinumab) or IL-12/IL-23 (e.g. ustekinumab).  The article does not provide further details on other exclusion criteria used in the study. |
| N_randomized | 676 | The total number of participants randomly assigned to a treatment group in the study was 676.  This is stated in the Results section:  "A total of 676 subjects were randomized. Among these, 675 subjects were included in the full analysis set for primary efficacy analyses (1 subject was excluded [informed consent was obtained the day after study-related procedure])." |
| N_randomized_per_group | Secukinumab: 337; Ustekinumab: 339 | The number of participants randomly assigned to each treatment group was:  - Secukinumab group: 337 participants  - Ustekinumab group: 339 participants  This is stated in Table 1, which shows the baseline characteristics for each group:  "Characteristic Secukinumab 300 mg (n = 337)  Ustekinumab (n = 339)"  So 337 participants were randomly assigned to the secukinumab group, and 339 participants were randomly assigned to the ustekinumab group. |
| N_analyzed | Secukinumab: 334; Ustekinumab: 335 | The number of participants analyzed in each treatment group was:  - Secukinumab group: 334  - Ustekinumab group: 335  This is stated when reporting the primary endpoint results in the Results section:   "Secukinumab (79.0%) was superior to ustekinumab (57.6%) as assessed by PASI 90 response at week 16 (P <.0001). The 100% improvement from baseline PASI score at week 16 was also significantly greater with secukinumab (44.3%) than ustekinumab (28.4%) (P < .0001)."  Where the sample sizes are noted as:  Secukinumab: 264/334 Ustekinumab: 193/335  So 334 participants from the secukinumab group and 335 from the ustekinumab group were analyzed for the primary endpoint. |
| Dose_route_frequency | Secukinumab: 300 mg, subcutaneous, weeks 0, 1, 2, 3, 4, and then every 4 weeks; Ustekinumab: 45 mg or 90 mg (for individuals with a weight of more than 100 kg), subcutaneous, weeks 0, 4, and then every 12 weeks | The dose, route of administration, and frequency for each treatment group were:  Secukinumab group: - Dose: 300 mg  - Route: Subcutaneous injection - Frequency: Weeks 0, 1, 2, 3, and every 4 weeks from week 4 to 48  Ustekinumab group:  - Dose: 45 mg for ≤100 kg body weight, 90 mg for >100 kg body weight - Route: Subcutaneous injection - Frequency: Weeks 0 and 4, then every 12 weeks from week 16 to 40  As stated in the Methods section:  "Eligible subjects were randomized 1:1 via an interactive response technology system to subcutaneous injection of secukinumab 300 mg or ustekinumab (dosing per label24: 45 mg for subjects #100 kg at baseline; 90 mg for subjects >100 kg at baseline)."   "Secukinumab was administered at baseline and weeks 1, 2, and 3, then every 4 weeks from week 4 to week 48; ustekinumab at baseline and week 4, then every 12 weeks from week 16 to week 40." |
| Primary outcome_name | PASI 90 | The primary outcome of the study was PASI 90 response at Week 16.   PASI 90 refers to ≥90% improvement in Psoriasis Area and Severity Index score from baseline.  This is stated in the Methods section:  "The primary objective was to demonstrate superiority of secukinumab versus ustekinumab with respect to PASI 90 response at week 16."  And reiterated in the Results:  "Secukinumab (79.0%) was superior to ustekinumab (57.6%) as assessed by PASI 90 response at week 16 (P <.0001)."  Therefore, the primary outcome was PASI 90 response at 16 weeks. |
| Primary outcome_estimate | Secukinumab: 79.0%; Ustekinumab: 57.6%, p<0.0001 | For the primary outcome of PASI 90 response at week 16:  Secukinumab group:  - 264/334 participants achieved PASI 90 response (79.0%)  Ustekinumab group: - 193/335 participants achieved PASI 90 response (57.6%)  P-value: <0.0001  This is reported in the Results section:  "Secukinumab (79.0%) was superior to ustekinumab (57.6%) as assessed by PASI 90 response at week 16 (P <.0001)."  Where the PASI 90 response is presented as a proportion of participants in each group achieving that endpoint. The p-value <0.0001 indicates the treatment difference was statistically significant. |

**Warren et al. 2021^11^**

| **Variable** | **Reference standard** | **Claude 2^9^** |
| --- | --- | --- |
| First author's last name | Warren | The first author's last name is Warren. |
| Trial registry number | NCT03478787 | The clinical trials registry number for this study is NCT03478787, as stated in the Methods section:  "IMMerge was a phase III, international, multicentre, randomized, open-label, efficacy–assessor-blinded, active-comparator study of up to 88 weeks’ total duration (Clinicaltrials.gov identifier: NCT03478787)." |
| Study name and acronym | IMMerge | The trial name is "Efficacy and safety of risankizumab vs. secukinumab in patients with moderate-to-severe plaque psoriasis (IMMerge)" and the acronym is IMMerge. This is stated in the title and abstract:  "Efficacy and safety of risankizumab vs. secukinumab in patients with moderate-to-severe plaque psoriasis (IMMerge): results from a phase III, randomized, open-label, efficacy–assessor-blinded clinical trial" |
| Study funder | AbbVie Inc | The study was funded by AbbVie Inc. This is stated in the Funding sources section:  "AbbVie Inc. funded this study, and participated in the study design, research, analysis, data collection, interpretation of data, reviewing and approval of the publication." |
| Mean age | Risankizumab: 47.3 (13.4); Secukinumab: 46.8 (14.9) | The average age of participants in each treatment group was:  - Risankizumab 150 mg: 47.3 years - Secukinumab 300 mg: 46.8 years  This is stated in the Patients and Methods section under Baseline demographics and disease characteristics:  "The mean ± SD patient age was 47.1 ± 14.1 years and 65.1% were male. At baseline, patients had a mean ± SD PASI of 19.9 ± 7.2, and 84.7% of patients had an sPGA score of 3."  And broken down by treatment group in Table 1:  "Age (years), mean ± SD  - Risankizumab 150 mg: 47.3 ± 13.4 - Secukinumab 300 mg: 46.8 ± 14.9" |
| Female participants | Risankizumab: 52 (31.7); Secukinumab: 62 (38.0) | Okay, let's go through this step-by-step:  - The study report states that overall, 65.1% of participants were male.   - There were 164 participants in the risankizumab group and 163 in the secukinumab group.  - In the risankizumab group, there were 112 males (68.3% of 164).  - So there must have been 164 - 112 = 52 females in the risankizumab group.  - 52 females is 52/164 = 31.7% of the risankizumab group (rounded to one decimal place).  - In the secukinumab group, there were 101 males (62.0% of 163).   - So there must have been 163 - 101 = 62 females in the secukinumab group.  - 62 females is 62/163 = 38.0% of the secukinumab group (rounded to one decimal place).  Therefore, the total count and percentage of females in each treatment group is:  - Risankizumab: 52 females (31.7%)  - Secukinumab: 62 females (38.0%) |
| Mean PASI score at baseline | Risankizumab: 19.8 (6.3); Secukinumab: 20.1 (8.1) | According to the baseline characteristics table:  - In the risankizumab group, the mean PASI score at baseline was 19.8 ± 6.3  - In the secukinumab group, the mean PASI score at baseline was 20.1 ± 8.1  So the mean PASI scores with standard deviations at baseline were:  - Risankizumab: 19.8 ± 6.3 - Secukinumab: 20.1 ± 8.1 |
| Mean duration of disease | Risankizumab: 18.6 (12.6); Secukinumab: 17.4 (13.2) | According to the baseline characteristics table:  - In the risankizumab group, the mean duration of plaque psoriasis was 18.6 ± 12.6 years - In the secukinumab group, the mean duration was 17.4 ± 13.2 years  Rounding the standard deviations to one decimal place:  - Risankizumab: 18.6 ± 12.6 years = 18.6 ± 12.6 years - Secukinumab: 17.4 ± 13.2 years = 17.4 ± 13.2 years  So the average number of years with standard deviation to one decimal place that participants had plaque psoriasis was:  - Risankizumab: 18.6 ± 12.6 years - Secukinumab: 17.4 ± 13.2 years |
| Inclusion criteria | Adults with moderate-to-severe chronic plaque psoriasis with or without psoriatic arthritis defined by PASI ≥ 12, sPGA ≥ 3, and BSA involvement ≥ 10%, candidate for systemic therapy. | The study inclusion criteria are described in the Patients section:  "This study recruited adult patients with a diagnosis of chronic, moderate-to-severe plaque psoriasis with or without psoriatic arthritis for at least 6 months before the baseline visit and who were candidates for systemic therapy including secukinumab. Patients had to demonstrate at least 10% body surface area covered with psoriasis plaques, a static Physician’s Global Assessment (sPGA) score of ≥ 3, and a Psoriasis Area and Severity Index (PASI) ≥ 12 at screening and baseline." |
| Exclusion criteria | History of erythrodermic psoriasis, generalized or localized pustular psoriasis, medication-induced psoriasis, new-onset guttate psoriasis, inflammatory bowel disease, chronic infections, active systemic infection, history of malignancy or previous exposure to risankizumab or secukinumab | The study exclusion criteria are listed in the Patients section:  "Patients were not eligible to participate if they had a history of erythrodermic psoriasis, generalized or localized pustular psoriasis, medication-induced or medication-exacerbated psoriasis, or new-onset guttate psoriasis (or any other active skin disease) that might interfere with the study assessments. Other exclusion criteria included a history of inflammatory bowel disease (based on warnings and precautions provided in the secukinumab prescribing information), chronic infections, active systemic infection (except common cold) during the last 2 weeks preceding the baseline visit, and history of malignancy, except for successfully treated nonmelanoma skin cancer or localized carcinoma in situ of the cervix, within the last 5 years. Previous exposure to risankizumab or secukinumab was not permitted." |
| N_randomized | 327 | The total number of participants randomly assigned to a treatment group is stated in the Results section:  "Of 409 patients screened, in total 327 from Canada, France, Germany, Italy, the Netherlands, Poland, Spain, the UK and the USA were randomized to risankizumab 150 mg (n = 164) or secukinumab 300 mg (n = 163)."  So the total number of participants randomly assigned was 327. |
| N_randomized_per_group | Risankizumab: 164; Secukinumab: 163 | The number of participants randomly assigned to each group is stated in the Results section:  "Of 409 patients screened, in total 327 from Canada, France, Germany, Italy, the Netherlands, Poland, Spain, the UK and the USA were randomized to risankizumab 150 mg (n = 164) or secukinumab 300 mg (n = 163)."  So the number of participants randomly assigned to each group was:  - Risankizumab 150 mg: 164 - Secukinumab 300 mg: 163 |
| N_analyzed | Risankizumab: 164; Secukinumab: 163 | The number of participants analyzed in each group is not directly stated, but can be inferred from the results reported.   In the Results section, it states the total number randomized to each group:  "Of 409 patients screened, in total 327 from Canada, France, Germany, Italy, the Netherlands, Poland, Spain, the UK and the USA were randomized to risankizumab 150 mg (n = 164) or secukinumab 300 mg (n = 163)."  Later in the Results section, when reporting efficacy outcomes at week 16, the number responding in each group is given:  "At week 16, 73·8% (n = 121) of patients randomized to risankizumab achieved PASI 90 compared with 65·6% (n = 107) of patients randomized to secukinumab."  Since 121 and 107 responders were reported for the risankizumab and secukinumab groups respectively, it can be inferred that the number analyzed was likely the same as those randomized, which was 164 in the risankizumab group and 163 in the secukinumab group. |
| Dose_route_frequency | Risankizumab: 150 mg, subcutaneous, weeks 0, 4, 16, 28, and 40, patients in France received additional doses at weeks 52 and 64; Secukinumab: 300 mg, subcutaneous, weeks 0, 1, 2, 3, and 4 then every 4 weeks for 48 weeks | According to the study design:  Risankizumab:   Dose: 150 mg (two 75 mg injections)  Route: Subcutaneous  Frequency: Weeks 0, 4, then every 12 weeks up to week 40  Additional doses: Patients in France received additional doses at weeks 52 and 64  Maximum length: 64 weeks  Secukinumab:   Dose: 300 mg (two 150 mg injections)  Route: Subcutaneous  Frequency: Weeks 0, 1, 2, 3, 4, then every 4 weeks up to week 48  Maximum length: 48 weeks |
| Primary outcome_name | PASI 90 | The primary outcomes of the study were:  1) The proportion of patients achieving ≥90% improvement in Psoriasis Area and Severity Index (PASI 90) at week 16.  2) The proportion of patients achieving PASI 90 at week 52.  This is stated in the Efficacy endpoints section:  "This study assessed two primary efficacy endpoints: (i) the proportion of patients achieving an improvement in PASI of at least 90% from baseline (PASI 90) at week 16 to assess the noninferiority of risankizumab vs. secukinumab (noninferiority margin of 12%), and (ii) the proportion of patients achieving PASI 90 at week 52 to assess the superiority of risankizumab vs. secukinumab." |
| Primary outcome_estimate | Primary outcomes At week 16 N (%) PASI 90 Risankizumab: 121 (73.8) Secukinumab: 107 (65.6) ARD (96.25% CI): 8.2% (–2.2 to 18.6) (within the 12% noninferiority margin) At week 52 N (%) PASI 90 Risankizumab: 142 (86.6) Secukinumab: 93 (57.1) ARD (95% CI): 29.8 (20.8 to 38.8), P < .001 | The primary outcomes were the proportions of patients achieving PASI 90 response at weeks 16 and 52.   At week 16: - Risankizumab: 73.8% (121/164 patients)  - Secukinumab: 65.6% (107/163 patients) - The adjusted difference was 8.2% (96.25% CI -2.2 to 18.6%), meeting non-inferiority   At week 52: - Risankizumab: 86.6% (142/164 patients) - Secukinumab: 57.1% (93/163 patients)  - The adjusted difference was 29.8% (95% CI 20.8 to 38.8%), p<0.001, demonstrating superiority  So risankizumab was non-inferior to secukinumab at week 16 and superior at week 52 for PASI 90 response based on the primary analysis. |
